# Spatial and single-cell transcriptomics on *SSPO* PTV mutant mice reveals SCO-spondin’s function in mood regulation through brain barriers

**DOI:** 10.1101/2024.04.09.24305433

**Authors:** Yue Gao, Jiahui Xu, Shuangyan Li, Wenjuan Yi, Liuyan Xiao, Fei Feng, Bin Zhang, Jiahui Li, Xinping Yang

## Abstract

The schizoaffective disorder presents mixed symptoms of schizophrenia and a mood disorder, posing an enormous challenge to its taxonomy and etiology. A recent hypothesis proposes independent genetic risk components from schizophrenia and affective disorders. By whole exome sequencing of a family with schizoaffective disorder, we identified four genes with protein-truncating variants (PTVs) that fit autosomal dominant inheritance model. Among these four genes, *SSPO* (encoding SCO-spondin) has exclusive expression in the subcommissural organ (SCO), a functionally highly intriguing organ in the brain, known to be potentially involved in CSF homeostasis. We generated a knock-in mouse model for the identified PTV of *SSPO*, and the mutant mice exhibited enhanced anxiety-like and anti-depression-like behaviors. We further performed multilayer transcriptomics analyses on the mouse brain, and found that the genes involved in stress response, anxiety, depression and bipolar disorder are dysregulated in the SCO and brain barriers, which have direct contact with CSF. Several cell-cell communication pathways involving brain barrier cells, including the WNT signaling pathways, are altered in the mutant mice. Our results demonstrate that SCO-spondin plays a key role in stress response and mood regulation through modulating brain barriers, and thus strongly suggest that the PTV of *SSPO* may contribute to the affective symptoms of schizoaffective disorder.

## Introduction

Schizoaffective disorder is a chronic mental health condition characterized primarily by symptoms of schizophrenia, such as hallucinations or delusions, and symptoms of a mood disorder, such as mania and depression ^1–3^. During its longitudinal course, patients with schizoaffective disorder can present with schizoaffective episodes, but also pure mood episodes similar to those of classic bipolar disorder and pure psychotic episodes, similar to those of typical schizophrenia ^4^, causing difficulties in its taxonomy and etiology ^5^. Although genetic epidemiologic studies support the existence of a strong genetic factor in schizoaffective disorder ^5,6^, the etiology of schizoaffective disorder remains unknown. The studies on the family, twin and adoption support the spectrum of symptoms as possibly being phenotypical expressions of genetic interforms of schizophrenia and affective psychoses ^5^. Because of the high genetic and phenotypic heterogeneity, the genetic linkage studies to search for disease genes of the affective disorder ^7–9^ and schizophrenia ^10–12^ have given controversial results. The heterogeneity is greater in schizoaffective disorder ^5^. Therefore, the identification of causal variants in a schizoaffective disorder family may need combination of whole exome sequencing (WES) and animal models. As this report will present, a protein-truncating variant (PTV) of *SSPO* identified in a pedigree with schizoaffective disorder using WES has been demonstrated to be involved in the mood-related symptoms using an animal model.

The *SSPO* is exclusively expressed in the subcommissural organ (SCO), an evolutionarily conserved brain structure located at the roof of the Sylvian aqueduct, beneath the posterior commissure. Although the SCO was first described a century ago and its functional importance was recognized as early as the 1950s ^13^, the function of the SCO remains a mystery. It has been known as a small gland consisting of ependymal and hypendymal cells ^13–15^, secreting the giant glycoprotein SCO-spondin into cerebrospinal fluid (CSF) which aggregates to form Reissner’s fiber (RF) ^14–17^. The earliest studies suggested its function in the regulation of electrolytes (such as Sodium and Potassium) and water balance in 1950s and 1960s ^14,18^. More recent studies suggest its roles in CSF circulation, homeostasis and clearance of certain compounds in CSF ^15,19,20^. It receives neural input, and is under neural control from multiple neural transmitter systems such as GABA, dopamine, noradrenaline and serotonin ^21–23^, suggesting that it may be involved in stress response and mood regulation, because these neurotransmitters control basic emotions and are implicated in affective disorders ^24^. Like other circumventricular organs (CVO), the SCO has axon terminals from the hypothalamus, secreting neurohormones, supporting that the SCO is functionally linked to the hypothalamus ^25^.

SCO-spondin, encoded by *SSPO*, which belongs to the group of spondins in the thrombospondin-1 family ^26^, is highly conserved in phylum chordates, and shows life-lasting expression ^27,28^. Some studies have shown that components from Reissner’s fiber or SCO-spondin-derived peptides inhibit angiogenesis ^29^, promote cell-to-cell interactions and neurite outgrowth of the neurons, and maintain neurite network in culture cells ^16,30,31^. In zebrafish, defects in SCO-spondin and Reissner’s fiber lead to neuroinflammatory response and spinal curvature through CSF ^32–34^. In human, it is reported that a genomic duplication spanning *ZNF467* and partial *SSPO* is identified in 39 patients out of 154 subjects in 68 bipolar families ^35^, and differential methylation of *SSPO* is identified in patients with irritable bowel syndrome ^36^. Using whole exome sequencing, *SSPO* has been found to be associated with irritable bowel syndrome ^37^, depression ^37^ and schizophrenia ^38^.

Here, in this study, we identified 4 protein-truncating variants (PTVs) in *SSPO*, *FGFRL1*, *ASPN* and *OR13C5*, in a schizoaffective family by whole exome sequencing (**Fig. 1a**). Considering the potential role of SCO-spondin and the SCO in mood regulation, we decided to verify the potential causal variant in *SSPO* and to understand the molecular pathogenesis using an animal model. We generated the *Sspo* PTV knock-in mouse model (**Fig. 1b**), and the mutant mice exhibited enhanced anxiety, anti-depression behaviors and increased pre-pulse inhibition (**Fig. 1b**). We carried out multilayer transcriptomics on the mouse brain, and found that the CSF-contacting brain regions, including blood-CSF barriers and CSF-brain barriers, are involved in mood-related behaviors of the mutant mice (**Fig. 1c**). Our results demonstrate that SCO-spondin plays a key role in stress response and mood regulation through modulating brain barriers (**Fig. 1d**), suggesting that SCO-spondin deficit caused by *SSPO* PTV may contribute to the affective symptoms of schizoaffective disorder.

**Fig. 1.**
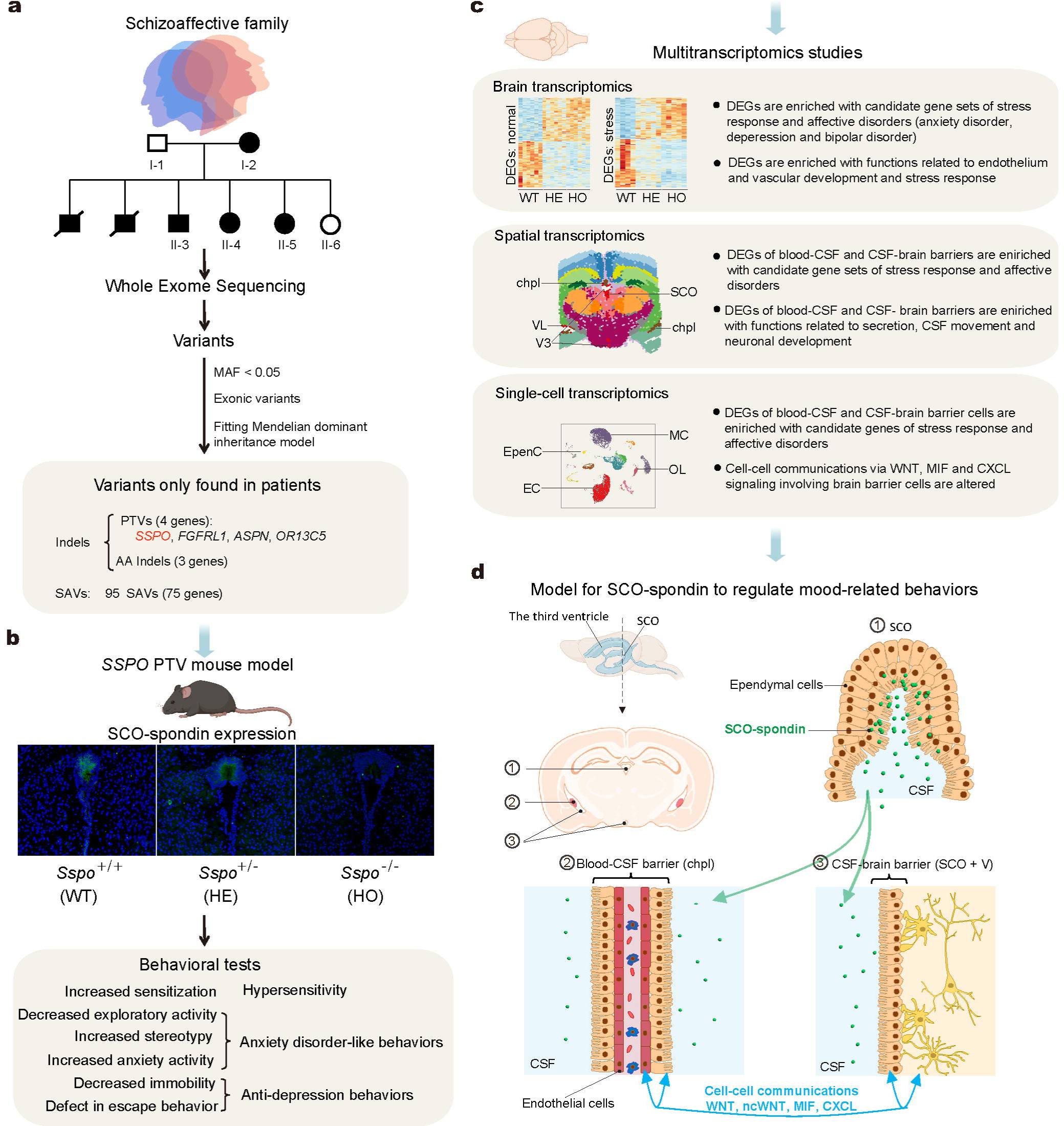
Overview. a. Identification of candidate causal variants in a family with schizoaffective disorder using whole exome sequencing (WES). b. The generation of SSPO-PTV knock-in mouse model and behavioral tests. Anxiety disorder-like and anti-depression behaviors detected in SSPO-PTV mice. c. Multitranscriptomics studies on the brain of the mouse model to illustrate the molecular mechanism underlying the affective symptom-like behaviors: (i) Bulk transcriptomics reveals that dysregulated genes are enriched with genes involved in affective disorders and endothelial functions; (ii) Spatial transcriptomics reveals that the CSF contacting brain regions, including the SCO, CSF-brain barrier, blood-CSF barrier, hypothalamus and olfactory area, are highly affected, and the dysregulated genes of these regions are enriched with genes involved in affective disorders, CSF secretion and movement; (iii) Single-cell transcriptomics reveals that the cell types that constitute blood-CSF and CSF-brain barriers are affected most, and the dysregulated genes of these cell types are enriched with genes involved in affective disorders, and cell-cell communication signaling pathways such as WNT, ncWNT, MIF and CXCL between these cell types and others are altered. d. The model for SCO-spondin to regulate mood through blood-CSF and CSF-brain barriers. The deficit of the SCO-spondin in the CSF can directly regulate brain barrier cells through binding SCO-spondin receptors, and induce dysfunction of barrier cells. The affected barrier cells may further alter cell-cell communications between themselves and other cell types. Brain regions: SCO: subcommissural organ; VL: lateral ventricles; V3: third ventricle; chpl: choroid plexus. Cell types: EpenC: ependymal cell; EC: endothelial cell; MC: microglial cell; OL: oligodendrocyte.

## Result

### Identification of a protein truncating variant (PTV) of *SSPO* in a schizoaffective family

We investigated a family with schizoaffective disorder (**Fig. 2a**), diagnosed based on the criteria from the BPRS (Brief Psychiatric Rating Scale) ^39^, Ham-D (or HDRS: Hamilton Depression Rating Scale) ^40^, PANSS (Positive and Negative Syndrome Scale) ^41^ and BRMS (Bech-Rafaelsen Mania Scale) ^42^. In this pedigree, the mother, three sons and two daughters are affected, and the father and another daughter are normal (**Fig. 2a**) (See Methods and **Table S1** for the clinical features). We carried out whole-exome sequencing on the blood samples of the individuals from this family and identified 43,662 exonic nonsynomymous SNVs in 14,652 genes and 2,340 exonic indels in 1,637 genes. Of these variants, 7 small indels in 7 genes and 95 nonsynonymous SNVs in 75 genes were found only in the affected individuals but not in the unaffected, fitting the mendelian autosomal dominant model with complete penetrance (**Fig. S1a**, **Table S1**). Of the 7 genes with indels, 4 have protein-truncating variants (PTVs), caused by a stop-gain or a frameshift (*SSPO*, *FGFRL1*, *ASPN and OR13C5*), and the other 3 genes have small indels of 1-2 amino acids. We speculated that these four genes with PTVs might be the best candidates for functional validation for establishing their roles in schizoaffective disorder. Of these four genes, *FGFRL1* and *OR13C5* have been reported to be associated with schizophrenia ^43,44^, while *SSPO* is potentially associated with bipolar disorder ^35^, irritable bowel syndrome ^37^, depressive disorder ^37^ and schizophrenia ^38^. In this study, we chose to generate *SSPO*-PTV mouse model, not only because of its association with mood disorders and schizophrenia, but also because of its exclusive expression in the SCO, a functionally unknown and intriguing organ in the brain. We verified the PTV mutation (p.Tyr3861>Stop) in *SSPO* of the affected members using PCR amplification followed by Sanger sequencing (**Fig. 2a, 2b**) before generating the mouse model.

**Fig. 2.**
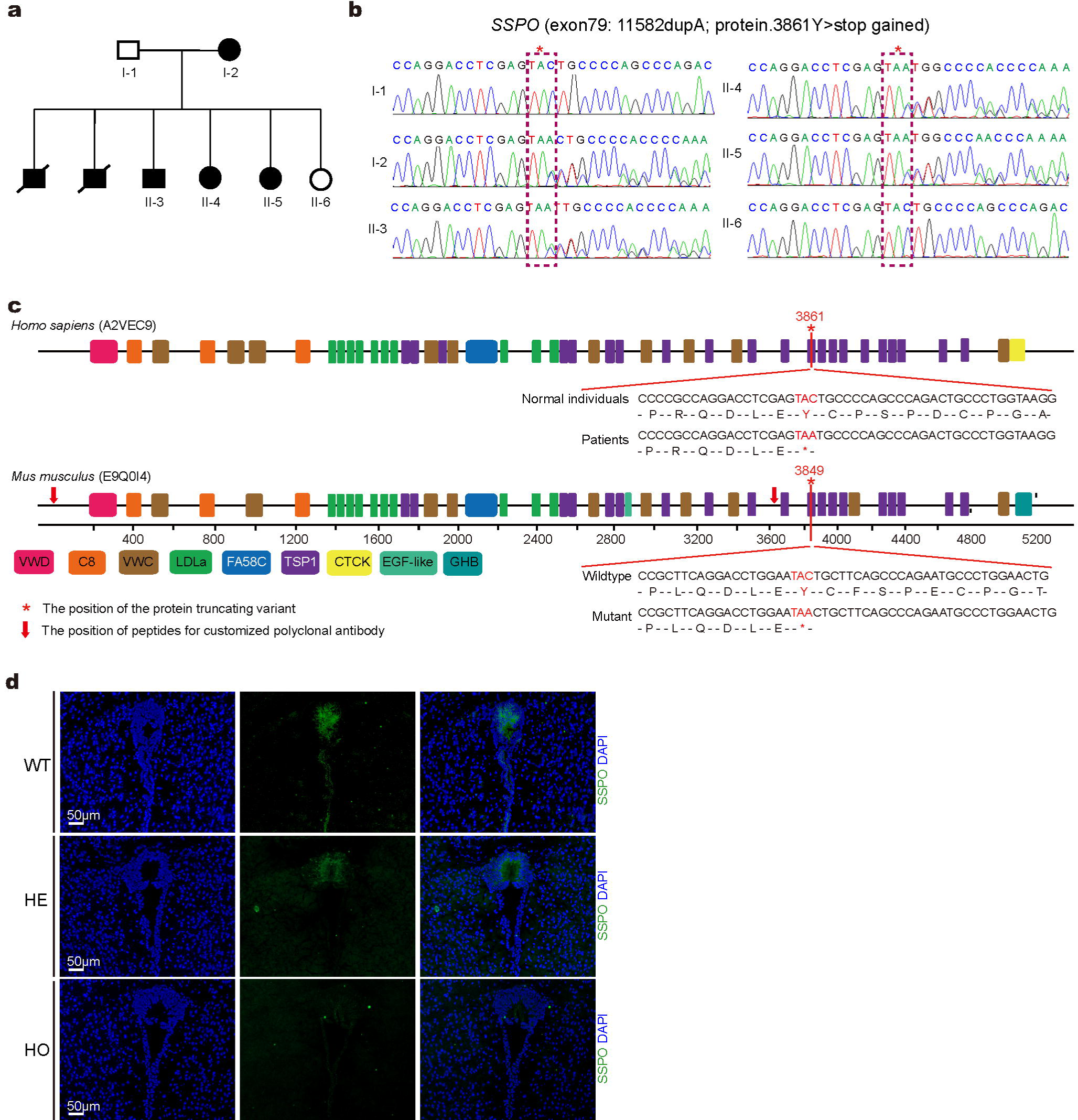
A protein-truncating variant (PTV) in *SSPO* in a family with schizoaffective disorder and the knock-in mouse model (See also Fig. S1). a. Pedigree of the schizoaffective family. The black boxes indicate the affected and white boxes indicate the unaffected. b. Verification of the “p.Tyr3861>Stop” mutation (chr7: 149818100: c.11582dupA) in *SSPO* identified by WES in the affected individuals using PCR amplification followed by Sanger sequencing. The red box shows the duplication of A at the mutation site. c. The protein domain structures of human and mouse SCO-spondins, showing the position of the stop-gain mutation in human *SSPO* and the corresponding position chr6: 48486792, c.11546dupA (tyr3849>Stop) in mouse *Sspo*. The red arrows show the positions of the peptides used for developing polyclonal antibodies. d. The immunofluorescence staining of SCO-spondin in the SCO of the knock-in mice using the custom antibody. The green fluorescence shows the SCO-spondin and the blue fluorescence shows the nuclei of cells in wild-type (WT), heterozygous (HE) and homozygous (HO) mice.

### The knock-in mouse model for the *SSPO*-PTV mutation

SCO-spondin has 76.26% sequence identity and a similar domain structure between human and mouse (**Fig. 2c**). The human *SSPO* PTV chr7: 149818100: c.11582dupA (p.Tyr3861>Stop) corresponds to chr6: 48486792, c.11546dupA (tyr3849>Stop) in the *Sspo* gene in mice (**Fig. 2c**). We generated the PTV in mice using CRISPR/Cas9 technology (**Fig. S1b,** see also Methods), and SCO-spondin was detected in the SCOs of the wild-type and heterozygous mutant mice, but not in the homozygous mutant mice (**Fig. 2d**), using immunofluorescence-labeled custom polyclonal antibodies (Sino Biological Inc., Beijing, China) against two synthesized peptides (HVEEEVTPRQEDLVPC, CPGPGIWSSWGPWEK) located in N-terminal part before the PTV site (**Fig. 2c**). The mutant mice exhibited normal growth, productivity, morphology and daily ambulation. Magnetic Resonance Imaging (MRI) was performed on the brain, and no abnormal brain structure was observed (**Fig. S1c**).

### Enhanced anxiety-like behaviors and anti-depression-like behaviors in the mutant mice

We carried out a repertoire of behavioral tests, including the open field, elevated plus maze, marble burying, tail suspension, learned helplessness and Y-maze tests, to investigate locomotion, exploratory activity, anxiety behaviors, repetitive behaviors, depression behaviors and working memory. We also evaluated pre-pulse inhibition (PPI), a measure of sensorimotor gating, because impairment in PPI had been reported to be associated with schizophrenia and several other psychotic disorders ^45^. The wild-type littermates were used as controls in these behavioral tests. One-way ANOVA was used to evaluate the effect of genotype in all these behavioral tests. Two-way ANOVA followed by Fisher’s LSD post hoc test for multiple comparisons was used in comparing the behaviors in the learned helplessness test and pre-pulse inhibition test. One-way ANOVA followed by Fisher’s LSD post hoc test for multiple comparisons was used to compare the behaviors in other tests.

#### Open field test

In the open field test, there was no significant effect of genotype on ambulation speed (One-way ANOVA, F (2, 28) = 0.5724, *p* = 0.5707, WT: n = 13; HE: n = 10; HO: n = 8), suggesting that the mutant mice have normal locomotion (**Fig. 3a-i**). The heterozygous mutant mice traveled significantly less distance in the center zone than the wild-type littermates (**Fig. 3a-ii**, *p =* 0.0164 for the heterozygous and *p =* 0.1766 for the homozygous). Consistently, the heterozygous spent less time in the center zone, while the homozygous showed a trend of less time in the center zone (**Fig. S2a**, *p =* 0.0439 for the heterozygous and *p =* 0.0888 for the homozygous). These results show that the mutant mice exhibit less exploring activity in center zone, which is usually interpreted as anxiety-like behaviors ^46^.

**Fig. 3.**
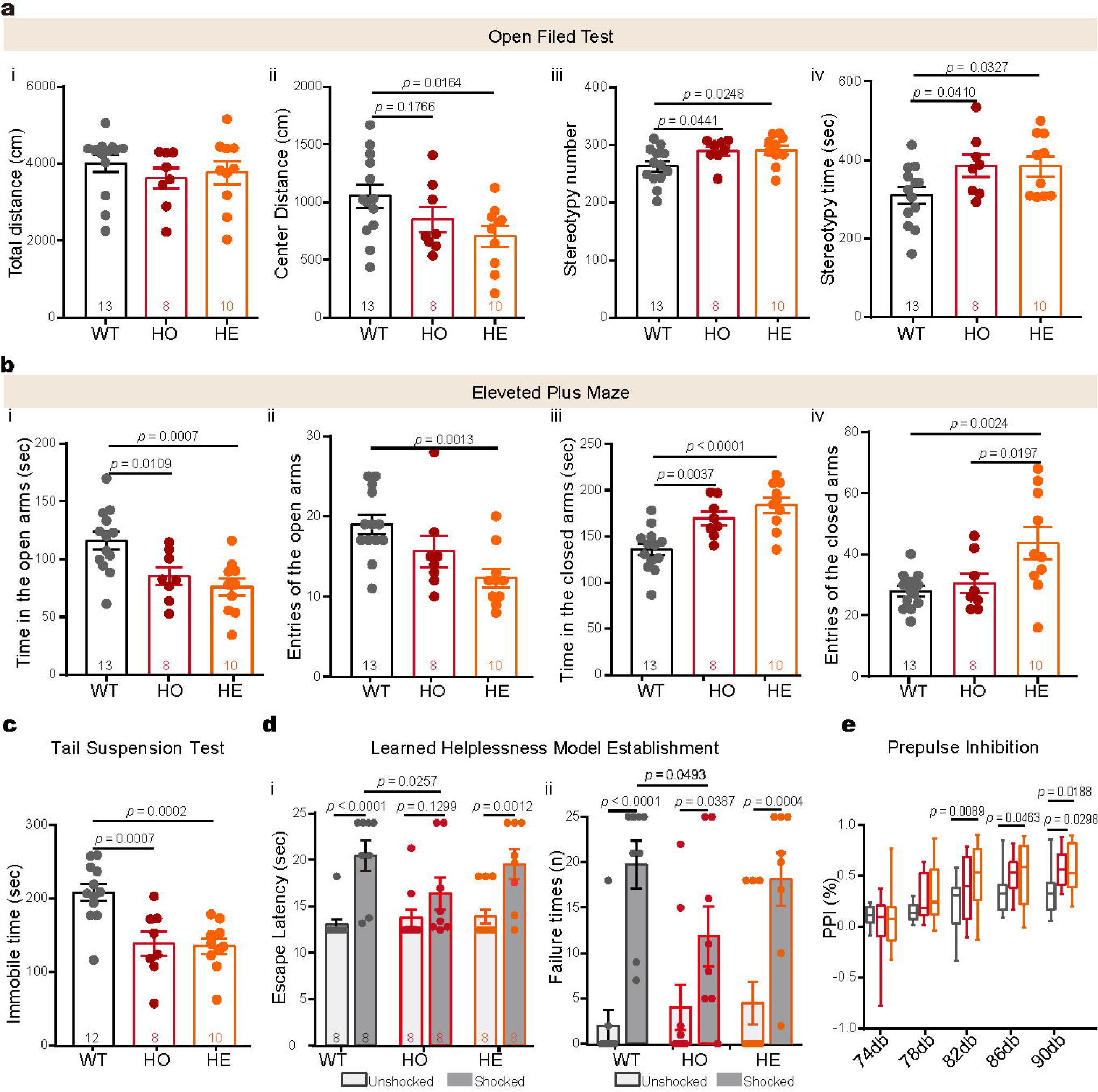
Anxiety behaviors and anti-depression behaviors in the *SSPO* mutant mice (See also Fig. S2). a. Open field test i. The total distance traveled. There was no significant effect of genotype on ambulation. ii. The distance traveled in the center zone. There was a significant effect of genotype on the distance traveled in the center zone. The heterozygous (HE) mutants displayed a lower percentage of distance traveled in the center zone than the wild-type (WT) mice, and the homozygous (HO) mutants showed a trend of lower percentage of distance traveled in the center zone than the wild-type mice. iii. The number of stereotyped behaviors in the open field test. The mutant (HE and HO) mice showed the increased number of stereotypies compared with the wild-type (WT) mice. iv. The duration of stereotyped behaviors in the open field test. The mutant (HE and HO) mice spent more time in stereotypic behaviors compared with the wild-type (WT) mice. b. Elevated plus maze. i. Duration in the open arms. Compared with the wild-type (WT) mice, both the heterozygous (HE) and the homozygous (HO) mutants spent significantly less time in the open arms. ii. The entries into the open arms. The heterozygous (HE) mutants displayed significantly fewer entries into the open arms compared with the wild-type (WT) mice. iii. The duration in the closed arms. Both the heterozygous (HE) and the homozygous (HO) mutants spent significantly more time in the closed arms than the wild-type (WT) mice. iv. The entries into closed arms. The heterozygous (HE) mutants entered the closed arms more frequently compared with the wild-type (WT) or the homozygous (HO) mice. c. The immobility in the tail suspension test. The mutant (HE and HO) mice displayed less immobility time compared with the wild-type (WT). d. Learned helplessness experiments i. Escape latency in learned helplessness experiments. Compared with the mice without 3 days of inescapable foot shocks, the mice with 3 days of foot-shock showed significant delayed escape in the wild-type (WT) and heterozygous (HE) mutant mice, but the homozygous (HO) showed no significant delayed escape. ii. Escape failure times in learned helplessness experiments. Compared with the mice without 3 days of inescapable foot shocks, the mice with 3 days of foot shock showed significant increased escape failure times in the wild-type (WT) and mutant (HO, HE) mice, but the homozygous showed relatively fewer failure times compared with the wild-type. e. Pre-pulse inhibition (PPI) tests. The genotypes showed a significant effect on PPI values at maximum sound stimulation. Compared with the wild-type (WT) littermates, the homozygous (HO) and heterozygous (HE) mutant mice, respectively, had significantly higher levels of pre-pulse inhibition (PPI %) at 82 db, 86 db and 90 db.

We also observed stereotypic behaviors in the open field test. There was a significant effect of genotype on stereotypic behaviors, including the effects on both the total number of stereotypies (**Fig. 3a-iii**, one-way ANOVA, F (2, 28) = 3.596, *p* = 0.0407) and the time spent in stereotypic activities (**Fig. 3a-iv**, one-way ANOVA, F (2, 28) = 3.429, *p =* 0.0466). Both the heterozygous and the homozygous displayed significantly more stereotypic behaviors (**Fig. 3a-iii**, *p =* 0.0248 for the heterozygous and *p =* 0.0441 for the homozygous) and spent more time in stereotypic behaviors compared to the wild-type littermates (**Fig. 3a-iv**, *p =* 0.0327 for the heterozygous and *p =* 0.0410 for the homozygous). Stereotypic behaviors refer to highly repetitive behaviors, freezing or other fear-related behaviors during the open field test. More stereotypic behaviors indicate an increased anxiety state ^47^.

#### Elevated plus maze test

In the elevated plus maze (EPM) test, the genotypes showed significant effects on the time spent in the open arms (one-way ANOVA, F (2, 28) = 8.052, *p =* 0.0017, WT: n = 13; HE: n = 10; HO: n = 8) and on the frequency entering the open arms (one-way ANOVA, F (2, 28) = 6.444, *p =* 0.0050) (**Fig. 3b**). Compared with the wild-type mice, both the heterozygous and homozygous mutant mice spent significantly less time in the open arms (**Fig. 3b-i**, *p =* 0.0007 for the heterozygous and *p =* 0.0109 for the homozygous), and the heterozygous entered the open arms significantly less frequently (**Fig. 3b-ii**, *p =* 0.0013 for the heterozygous and *p =* 0.1024 for the homozygous). The genotypes also had significant effects on the time spent in the closed arms (one-way ANOVA, F (2, 28) = 12.46, *p =* 0.0001) and on the frequency entering the closed arms (one-way ANOVA, F (2, 28) = 5.998, *p* = 0.0068). Both the heterozygous and homozygous mutant mice spent significantly more time in the closed arms (**Fig. 3b-iii**, *p* < 0.0001 for the heterozygous and *p =* 0.0037 for the homozygous), and the heterozygous mice entered the closed arms more frequently compared with the wild-type and homozygous mice respectively (**Fig. 3b-iv**, *p =* 0.0197 between the heterozygous and the homozygous, and *p =* 0.0024 between the heterozygous and the wild type). The total distance (**Fig. S2b**, one-way ANOVA, F (2, 28) = 0.346, *p =* 0.7105) and the number of transitions in all arms (**Fig. S2c**, one-way ANOVA, F (2, 28) = 2.17, *p* = 0.1330) were not significantly different between the three genotype groups; therefore, the less time and frequency exploring the open arms were not due to altered locomotor activity. These results further validate the increased anxiety in the mutant mice.

#### Marble burying test

In the marble burying test, the genotypes showed significant effect on the number of buried marbles (one-way ANOVA, F (2, 28) = 3.369, *p =* 0.0489, WT: n = 13; HE: n = 10; HO: n = 8). The heterozygous mutant mice showed a trend of burying more marbles compared with the wild-type mice (**Fig. S2d**, *p* = 0.0686 for between the heterozygous and the wild type). The marble burying behavior can be interpreted as anxiety-related or an obsessive/compulsive-like behavior ^48^.

#### Tail suspension test

In the tail suspension test, the genotypes showed significant effect on the immobility levels (one-way ANOVA, F (2, 27) = 11.67, *p =* 0.0002, WT: n = 12; HE: n = 10; HO: n = 8). Both the heterozygous and homozygous mutant mice displayed lower immobility levels compared with the wild-type animals (**Fig. 3c**, *p =* 0.0002 for the heterozygous and *p =* 0.0007 for the homozygous). This behavioral test measures depression-related resignation and is often used in screening for potential antidepressants ^49^. These results demonstrate consistent anti-depression behaviors in both the homozygous and heterozygous mutant mice.

#### Learned helplessness test

Since immobility in the tail suspension test is a direct reaction to the test itself rather than a persistent state of a depression model ^50^, we further performed the learned helplessness test to evaluate the ability to cope with an aversive situation as a measure of depressive states of mice ^51^. The mice were exposed to a series of inescapable foot shocks (0.3 mA, 100 times per day) for 3 days. After 3 days of foot shocks, the mice showed a significant effect of shocks on escape latency (**Fig. 3d-i**, two-way ANOVA, F (1, 50) = 29.04, *p* < 0.0001, shock group: WT: n = 8; HE: n = 8; HO: n = 8, control group: WT: n = 8; HE: n = 8; HO: n = 8). The mice exhibited a trend of (but not significant) effect of genotypes on the escape latency (Two-way ANOVA, genotype × shock interaction, F (2, 50) = 2.049, *p* = 0.1396). Compared with the mice without 3 days of inescapable foot shocks, the mice with 3 days of foot shocks showed a significant delay in escape in the wild-type and heterozygous mutant mice (**Fig. 3d-i**, *p* < 0.0001 for wild-type mice, *p =* 0.0012 for heterozygous mutant mice), but the homozygous showed no significant delay in escape (**Fig. 3d-i**, *p =* 0.1299 between test group and control group). The homozygous mice showed less escape latency compared with the wild-type mice (**Fig. 3d-i**, *p* = 0.0257).

The mice showed a significant effect of pretreament with 3 days of inescapable foot shocks on the number of failed escapes in the test (**Fig. 3D-ii**, two-way ANOVA, F (1, 50) = 38.28, *p* < 0.0001). Compared with the mice without 3 days of inescapable foot shocks, the mice with 3 days of foot shocks showed a significant increase in failed escapes in both the wild-type and mutant mice (**Fig. 3d-ii**, *p* < 0.0001 for the wild-type, *p =* 0.0004 for the heterozygous, *p =* 0.0387 for the homozygous). The mice exhibited a trend of genotypic effect on the failed escapes (Two-way ANOVA, genotype × shock interaction, F (2, 50) =1.789, *p =*0.1776). For the mice with 3 days of inescapable foot shocks, the homozygous showed fewer failed escapes compared with the wild-type (**Fig. 3d-ii**, *p =* 0.0493). These results show that the homozygous mutant mice have less learned helplessness, suggesting that the homozygous (not the heterozygous) mutant mice may have attenuated escape-failure-induced depressive state. The learned helplessness test shows that the homozygous mutant genotype has an anti-depression effect.

#### Y maze test

We measured spontaneous alternation in the Y-maze test, and detected no differences between the mutant and wild-type mice (**Fig. S2e-f,** WT: n = 13; HE: n = 10; HO: n = 8), indicating that the working memory was not impaired in the mutant mice.

#### Pre-pulse inhibition test

The pre-pulse inhibition (PPI) test is designed to assess the sensorimotor gating ability of the brain. The mice showed significant genotypic effect on pre-pulse inhibition (PPI%) values at sound stimulation (Two-way ANOVA, genotype × stimulation interaction, F (8, 112) = 2.899, *p* = 0.0056, WT: n = 13; HE: n = 10; HO: n = 8). The heterozygous and homozygous mutant mice had significantly higher levels of PPI% than the wild-type mice respectively (**Fig. 3e**, *p =* 0.0089 for the heterozygous mice at 82 db; *p =* 0.0463 for the heterozygous mice at 86 db; *p =* 0.0188 for the heterozygous mice and *p =* 0.0298 for the homozygous mice at 90 db), while the genotypes showed no significant differences in the stimulation of white noise (**Fig. S2g**). These results are consistent with increased anxiety observed in the mutant mice, because pre-pulse inhibition is reported to be increased in mice with anxiety-like behaviors ^52^.

These behavioral tests demonstrate that both the homozygous and heterozygous mutant mice have increased anxiety behaviors, less depressive behaviors and increased PPI% (See behavioral test summary in **Fig. S2h**), which can be interpreted as similar phenotypes to the increased anxiety and irritability in schizoaffective disorder ^53^. These results suggest that the *SSPO* PTV may contribute to the affective symptoms observed in the patients with schizoaffective disorder.

### Genes involved in endothelial functions, stress response and affective disorders are dysregulated in the brain of the mutant mice

A key motivation for generating the *SSPO*-PTV knock-in mouse model was to utilize a multi-omics approach to exploring the potential role of *SSPO* in schizoaffective disorder in human. Therefore, we further carried out bulk, spatial and single-cell transcriptomics on the brain to search for dysregulated genes and molecular pathways in different brain regions and cell types. For bulk transcriptomics, we performed two batches of RNA-seq on the whole brains of mice. The first batch was done on the mice (male, n = 6 for each genotype, age = 11w) taken from normal home cage condition, without undergoing the stress of behavioral tests, and the second batch on the mice (male, n = 5 for each genotype, age = 11w) which were sacrificed 3 hours after the acute stress of the tail suspension test. The samples of the three different genotypes were clustered in three separated groups in the principal component analysis (PCA) on the RNA-seq data of the mice under normal and stress conditions, respectively (**Fig. S3a-b**). The differential expression analysis was done using DeSeq2.

For the mice without exposure to stress condition, we identified 982 DEGs (493 up-regulated and 489 down-regulated) in the homozygous mutant mice and 622 (300 up-regulated and 322 down-regulated) in the heterozygous mutant mice compared to the wild-type mice (*p* < 0.01 and |Fold Change| ≥ 1.3, **Fig. 4a**, **Table S2**). The heterozygous have a similar gene expression pattern with the homozygous, based on the genome-wide threshold-free comparison using rank-rank hypergeometric pair-wise comparison ^54^ (**Fig. 4b**). The expression changes of the DEGs are well correlated between the two mutant genotypes (**Fig. 4c**, r = 0.91, *p* < 2.2e-16, Pearson’s product-moment correlation). Of the 622 DEGs found in the heterozygous, 62.06% (386 / 622) are included in the DEGs of the homozygous. These 386 common DEGs have larger fold changes in the homozygous than in the heterozygous (**Fig. 4c**), suggesting that the homozygous genotype may have greater genetic effects on brain gene expression than the heterozygous genotype.

**Fig. 4.**
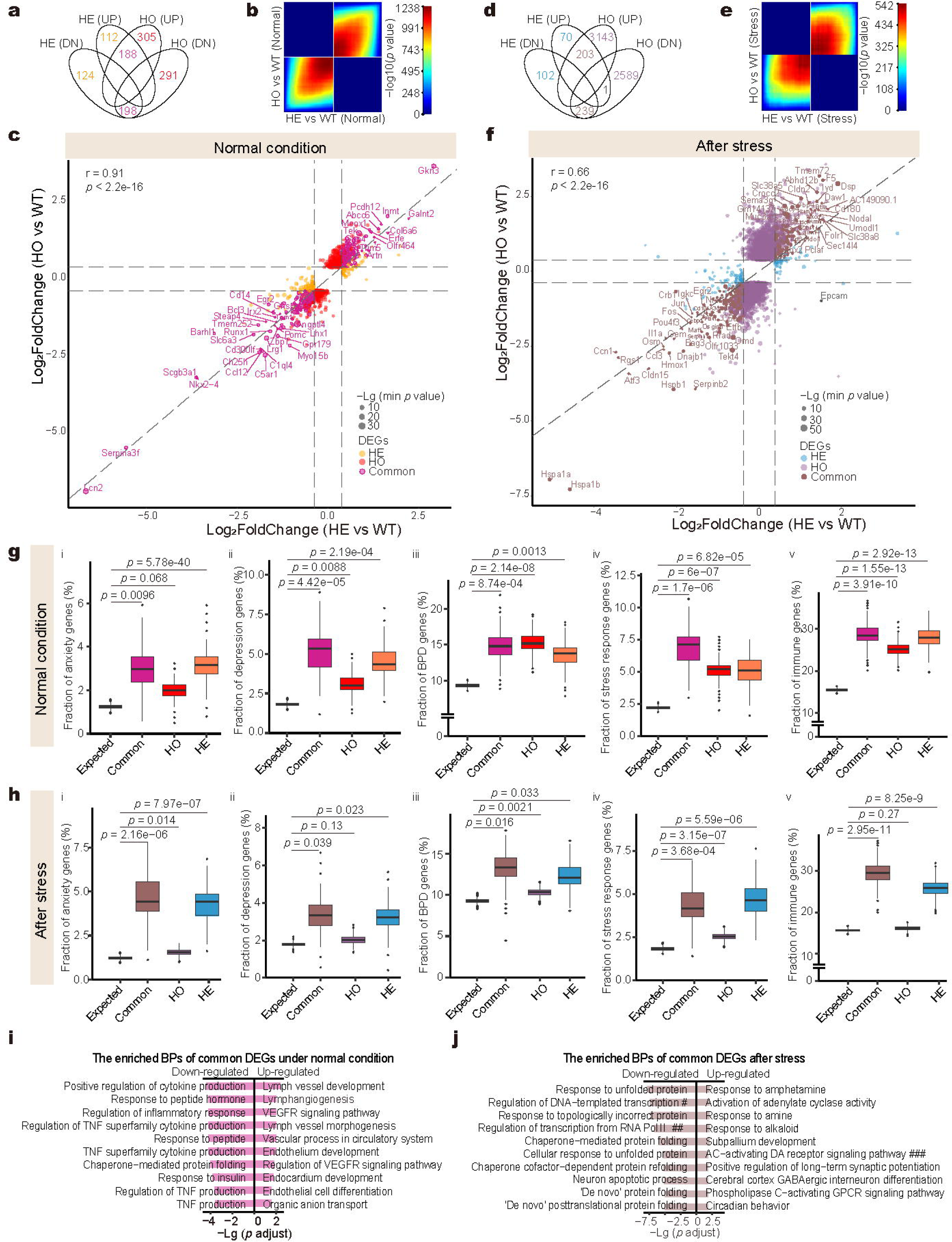
Genes involved in endothelium functions, stress response, anxiety and depression are dysregulated in the brain of the mutant mice (See also Fig. S3). a. The Venn diagrams of the DEGs in the brain of mice under normal condition: the homozygous (HO) and heterozygous (HO) mice compared with the wild-type (WT) mice (*p* < 0.01 and |FC|≥1.3). UP: up-regulated; DN: down-regulated. b. The threshold-free rank-rank hypergeometric (RRHO) pairwise comparison of the differential expression signatures of the heterozygous (HE) and homozygous (HO) mutants under normal condition. Points in the RRHO2 plot are colored by the *p* value of hypergeometric tests measuring the significance of the overlap of the gene lists up to that point. Genes from each signature are ordered from the most up-regulated to the most down-regulated, with the most up-regulated genes in the lower left corner. c. Scatter plot of the common DEGs of the heterozygous (HE) and homozygous (HO) mutants under normal condition. The Y-axis shows log2|fold change| of (HO vs WT), X-axis shows log2|fold change| of (HE vs WT). Orange, red and magenta represent the DEGs of the HE and HO mutants and common DEGs, respectively. The sizes of dots represent the values of –lg10 (*p* value HE vs WT) or –lg (*p* value HO vs WT). d. Venn diagram of the DEGs in mice under stress condition: the heterozygous (HE) and the homozygous (HO) mutant mice compared with the wild-type mice (*p* < 0.01 and |FC| ≥ 1.3). e. Genome-wide threshold-free hypergeometric (RRHO) pairwise comparison of the differential expression signatures between the heterozygous (HE) and homozygous (HO) mutants under stress condition. Points in the RRHO2 plot are colored by the *p* value of hypergeometric tests measuring the significance of the overlap of the gene lists up to that point. Genes from each signature are ordered from the most up-regulated to the most down-regulated, with the most up-regulated genes of each signature in the lower left corner. f. Scatter plot of the common DEGs of the heterozygous (HE) and homozygous (HO) mutants under stress condition. The Y-axis shows log2(fold change) of the differential expression values (HO vs WT), X-axis shows log2(fold change) of the differential expression values (HE vs WT). Blue, purple and brown represent the DEGs of the heterozygous (HE) and homozygous (HO) mutants, and the common DEGs, respectively. The sizes of dots represent the values of –lg (*p* value HE vs WT) or –lg (*p* value HO vs WT). g-h. The box and whisker graph of the fractions of the candidate gene sets of anxiety (g-i, h-i), depression (g-ii, h-ii), bipolar disorder (g-iii, h-iii), stress response (g-iv, h-iv) and immunity (g-v, h-v) in the DEGs of the homozygous (HO) and heterozygous (HE) mutants and the common DEGs, compared with all expressed genes in the brain. i. The enriched biological processes (BPs) of the common DEGs between the heterozygous (HE) and the homozygous (HO) under normal condition: left for the down-regulated DEGs and right for the up-regulated DEGs. j. The enriched biological processes (BPs) of the common DEGs between the heterozygous (HE) and the homozygous (HO) under stress condition: left for the down-regulated DEGs and right for the up-regulated DEGs. #: regulation of DNA−templated transcription in response to stress; ##: regulation of transcription from RNA polymerase II promoter in response to stress; ###: adenylate cyclase−activating dopamine receptor signaling pathway.

For the mice exposed to the acute stress of the tail suspension test, we identified 6,175 DEGs (3,346 up-regulated and 2,829 down-regulated) in the homozygous and 615 (274 up-regulated and 341 down-regulated) in the heterozygous, compared to the wild-type respectively (*p* < 0.01 and |Fold Change| ≥ 1.3, **Fig. 4d**, **Table S2**). Although the heterozygous and the homozygous under stress condition had a large difference in the number of DEGs, they had a similar gene expression pattern (**Fig. 4e**) based on the genome-wide threshold-free comparison ^54^, and the expression changes of the DEGs showed good correlation between the heterozygous and the homozygous (**Fig. 4f**, r = 0.66, *p* < 2.2e-16, Pearson’s product-moment correlation). Of the 615 DEGs found in the heterozygous, 72.03% (443/615) are included in the DEGs of the homozygous. These common 443 DEGs have larger fold changes in the homozygous than in the heterozygous, with only one gene showing a reverse change (**Fig. 4f**, **Table S2**). These results suggest that the homozygous genotype may have much greater genetic effects on brain gene expression than the heterozygous genotype and that the common dysregulated genes under acute stress condition may contribute to the similar stress response behaviors of the homozygous and heterozygous mutant mice. The transcription level of *Sspo* was significantly decreased in the mutant mice under normal condition (**Fig. S3c**). However, after the acute stress, the mRNA expression level increased 3.53-fold in the homozygous mice and 1.62-fold in the heterozygous mice compared with the wild-type mice (**Fig. S3c**), suggesting that SCO-spondin might be involved in stress response. We also looked at the expression of an ependymal marker gene, *Foxj1,* which is known to be specifically expressed in the ependymal cells of the SCO and the ventricles. The expression of *Foxj1* was also decreased in the mutants under normal condition, suggesting that ependymal cells may be affected in the mutant mice. However, the expression of *Foxj1* increased under stress condition (**Fig. S3c**), suggesting that ependymal cells may be involved in stress response.

To understand the behavioral phenotypes of the mutant mice and their potential indications in mental disorders, we calculated the enrichment of different candidate gene sets of mental disorders in the DEGs. The DEGs of the mutant genotypes under both normal and stress conditions were enriched with the candidate gene sets of anxiety (**Fig. 4g-i** and **h-i**), depression (**Fig. 4g-ii** and **h-ii**), bipolar disorder (BPD) (**Fig. 4g-iii** and **h-iii**), stress response (**Fig. 4g-iv** and **h-iv**) and immunity (**Fig. 4g-v** and **h-v**). However, these DEG sets are not enriched with the candidate gene sets of either schizophrenia (SCZ) (**Fig. S3d-i** and **e-i**) or autism spectrum disorder (ASD) (**Fig. S3d-ii** and **e-ii**). These results suggest that the SSPO PTV may be involved only in affective symptoms of schizoaffective disorder, not in psychotic symptoms.

We explored the functions of the common DEGs of the heterozygous and homozygous under normal and stress conditions, respectively, by the enrichment of biological processes of Gene Ontology (GO). For the 386 common DEGs of the mutant mice under normal condition, the up-regulated genes are mainly enriched with functions in vascular development and endothelial proliferation (**Fig. 4i**), while the down-regulated genes are enriched with functions in regulation of cytokine production, inflammatory response, hormone transportation and secretion (**Fig. 4i**). For the 443 common DEGs under stress condition, the up-regulated genes are enriched with functions such as response to amphetamine, adenylate cyclase-activating receptor related signaling pathways and positive regulation of long-term potential (**Fig. 4j**), which are reported to be involved in stress response ^55–57^, while the down-regulated genes are enriched with functions such as response to unfolded protein, transcription regulation in stress response and neuron apoptotic process (**Fig. 4j**), which are also reported to be involved in stress response ^58,59^. These results suggest that the blood-brain barriers, immunity and the ability to handle stress may be affected in the mutant mice.

### *SSPO*-PTV exerts effects on transcription of genes in brain regions associated with affective disorders

To determine the potential effects of the *SSPO*-PTV on different brain regions, we carried out spatial transcriptome sequencing using 10× Genomics Visium Spatial Gene Expression assay. The fresh-frozen brains from mice under normal condition were embedded in optimal cutting temperature (OCT) compound and cryosectioned to obtain coronal sections. For each brain slice, about 3,500 to 3,900 spots (on average, 3,941 for the wild-type, 3,611 for the heterozygous and 3,459 for the homozygous) were profiled, with 546 to 9,557 genes identified in each spot. The spots of all three slices were clustered into 16 spatial clusters based on the gene expression similarity using Seurat (**Fig. S4a**), and the anatomical annotation of the clusters was done by comparing their positions on the slice with the brain map of Allen Brain Atlas (**Fig. 5a** and **Fig. S4a**). The average detected genes of each cluster ranged from 3,854 to 7,405 genes (**Fig. 5b** and **Fig. S4b**, **Table S3**).

**Fig. 5.**
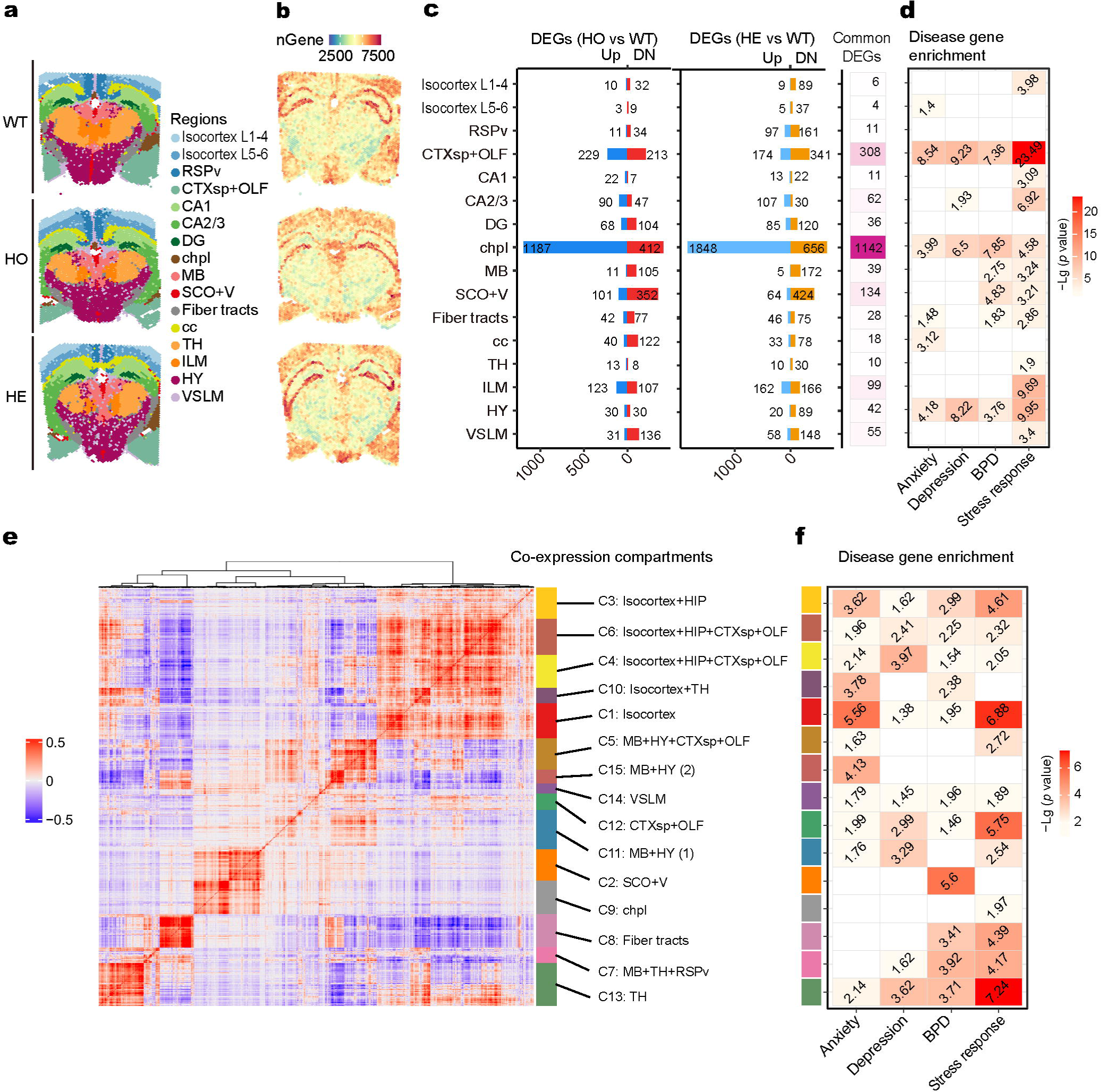
The altered spatial gene expression patterns associated with affective disorders (See also Fig. S4) a. The 16 spatial clusters of the spots based on gene expression similarity using Seurat. The 16 spatial clusters were annotated according to the brain map of Allen Brain Atlas. Isocortex L1-4: isocortex layer 1-4; Isocortex L5-6: isocortex layer 5-6; RSPv: retrosplenial area, ventral part; CTXsp: cortical subplate; OLF: olfactory area; CA1: hippocampus subfield CA1; CA2/3: hippocampus subfields CA2-3; DG: dentate gyrus; chpl: choroid plexus; MB: midbrain; SCO: subcommissural organ; V3: the third ventricle; Fiber tracts; cc: corpus callosum; TH: thalamus; ILM: intralaminar nuclei of the dorsal thalamus; HY: hypothalamus; VSLM: vascular leptomeninges. b. The distribution of gene number identified in the spots. The scaled colors represent the numbers of genes in each spot. c. The DEGs of each spatial cluster. The differential expression was calculated by comparing expression values of spots in each region between the mutant mice and the wild-type mice. Bar plots show the number of DEGs in the homozygous (HO) (left) and the heterozygous (HE) (right). Red and blue bars show the up-regulated and the down-regulated genes, respectively. The common DEGs are listed on the right. d. The enrichment of the candidate gene sets in the common DEGs of each spatial cluster. The overrepresentation of candidate gene sets of anxiety, depression, bipolar disorder and stress response was calculated for the common DEGs of each region. The heatmap shows the significance values of overrepresentation of the gene sets in the DEGs in each region. The scaled colors represent the enrichment scores. The numbers in the squares are the –lg (*p*-value). One-sided Fisher’s exact test was used for the enrichment analyses. e. Heatmap of gene co-expression matrix, showing the 15 spatial co-expression compartments. Co-expression was calculated for each gene in all spots. The top 2,000 co-expressed feature genes were further used for clustering the spots to obtain co-expression compartments. The brain regions in each compartment were annotated according to the brain map of the Allen Brain Atlas. The color bar on the right shows the spatial co-expression clusters (compartments) matching combinations of brain regions based on the brain map of the Allen Brain Atlas. The scale color bar at the right bottom corner shows the co-expression scores. f. The disease gene enrichment of the 15 spatial co-expression compartments. The overrepresentation of the candidate gene sets of anxiety, depression, bipolar disorder and stress response was calculated for the feature genes in each compartment. The heatmap shows the significance values of overrepresentation of the gene sets in the spatial co-expression compartments. One-sided Fisher’s exact test was used for the analyses of enrichments. The numbers in the squares represent the –lg (*p*-value).

We further performed gene differential expression analysis on each of the 16 spatial clusters between the mutant and wild-type mice. The most affected spatial clusters include “choroid plexus (chpl)” (HO: 1,599 DEGs; HE: 2,504 DEGs; Common: 1142 DEGs), “cortical subplate + olfactory area (CTXsp + OLF)” (HO: 442 DEGs; HE: 515 DEGs; Common: 308 DEGs), “SCO + ventricles (SCO + V)” (HO: 453 DEGs; HE: 488 DEGs; Common: 134 DGEs) and “intralaminar nuclei of the dorsal thalamus (ILM)” (HO: 230 DEGs; HE: 328 DEGs; Common: 89 DEGs) (**Fig. 5c**, **Table S3**). To find out if the functions of the dysregulated genes in the spatial clusters were with association to mood-related phenotypes, we calculated the enrichment of the common DEGs with the candidate gene sets of stress response and affective disorders, such as anxiety, depression and bipolar disorder, respectively (**Fig. 5d, Table S3**). DEGs of the spatial clusters “choroid plexus (chpl)”, “cortical subplate + olfactory area (CTXsp + OLF)” and “hypothalamus (HY)” are enriched with the candidate gene sets for stress response and three affective disorders respectively, while DEGs of the cluster “SCO + ventricles (SCO + V)” are enriched with candidate gene sets for stress response and bipolar disorder. Interestingly, the SCO, ventricles and choroid plexus have direct contact with CSF, while the hypothalamus and olfactory area have CSF-contacting neurons ^60,61^. CSF-contacting neurons serve as chemoreceptors and mechanosensors for sensing the chemical and mechanical changes in the CSF ^62^. These results show that the expression of the genes involved in stress response and affective disorders is disrupted in the CSF-contacting brain regions of the *SSPO*-PTV mutant mice, suggesting that the SCO-spondin may regulate stress response and mood-related behaviors through these CSF-contacting brain regions.

We wondered, apart from the highly affected brain regions in the *SSPO*-PTV mutant mice, if there were other regions participating in mood regulation and stress response. To explore the functional association between brain regions and affective phenotypic behaviors, we used Giotto (see Methods) to identify the co-expression compartments with coherent spatial expression patterns. The top 2,000 spatial expression values were used to calculate pairwise distances between genes, and 15 co-expression compartments were identified, each covering one or more of the above 16 spatial clusters (**Fig. 5e, Fig. S4c**). We calculated the enrichment of the candidate gene sets of anxiety, depression, bipolar disorder and stress response in each of these compartments (C1-15). Six compartments (C1, C10, C15, C3, C4, C13), covering 8 brain regions including isocortex, thalamus, hypothalamus, hippocampus, midbrain, cortical subplate and olfactory area, are significantly enriched with anxiety candidate genes (**Fig. 5f**, *p* < 0.01). Five compartments (C4, C13, C11, C12, C6), covering the same 8 brain regions, are significantly enriched with depression candidate genes (**Fig. 5f**, *p* < 0.01). These results are consistent with previous reports that these brain regions are found to be associated with human anxiety and depression by PET/MRI ^63,64^. Seven compartments (C2, C3, C6, C7, C8, C10, C13), covering the brain regions SCO, ventricles, isocortex, hippocampus, cortical subplate, olfactory area, thalamus, and fiber tracts, are significantly enriched with bipolar disorder candidate genes (**Fig. 5f**, *p* < 0.01). The enriched brain regions for bipolar disorder mostly overlap with those for anxiety and depression. The compartment 2 (C2), which contains the SCO and the ventricles, is exclusively enriched with bipolar disorder candidate genes (**Fig. 5f**). Nine compartments (C3, C4, C5, C6, C7, C8, C11, C12, C13), covering brain regions isocortex, hippocampus, cortical subplate, olfactory area, midbrain, hypothalamus, thalamus and fiber tracts, are significantly enriched with stress response genes (**Fig. 5f***, p* < 0.01). These brain regions mostly overlap with those enriched regions for anxiety, depression and bipolar disorders, suggesting a converged molecular mechanism underlying the stress response and affective disorders. The identified spatial clusters “SCO + ventricles (SCO + V)”, “choroid plexus (chpl)”, “cortex subplate + olfactory (CTXsp + OLF)” and “hypothalamus (HYP)” in the *SSPO*-PTV mutant mice (**Fig. 5d**) are included in the 15 co-expression compartments that show enrichment with candidate gene sets of stress response or affective disorders (**Fig. 5f**). As described above, the SCO, ventricles and choroid plexus have direct contact with CSF, while the hypothalamus and olfactory area have CSF-contacting neurons ^60,61^. Therefore, these spatial profiling results strongly support that, among the brain regions involved in stress response and mood regulation, the *SSPO* PTV mainly affects the CSF-brain barriers, CSF-blood barriers and the brain regions with cells contacting CSF.

### *SSPO*-PTV induces differential expression of genes involved in the secretion, flow and homeostasis of CSF

The spots in the SCO and ventricles are clustered together based on gene expression similarity (**Fig. S4a**). To identify genes with region-specific expression, we further compared the gene expression between the spots within the cluster “SCO + ventricles (SCO + V)” and the spots in other brain regions on the three slices, identifying 86 genes preferentially expressed in the SCO and the ventricles (**Fig. 6a** and **Fig. S5a**, **Table S3**). Similarly, we identified 180 genes preferentially expressed in the cluster “choroid plexus (chpl)” (**Fig. S5b**, **Table S3**). As expected, the traditional ependymal marker *Foxj1* are specifically expressed in the clusters “SCO + ventricles (SCO + V)” and “choroid plexus (chpl)”. We mapped the expression data of *Foxj1* and *Sspo* on the slices, finding that *Foxj1* was detected in the SCO, ventricles (V) and choroid plexus, while *Sspo* was exclusively detected in the SCO (**Fig. 6b**).

**Fig. 6.**
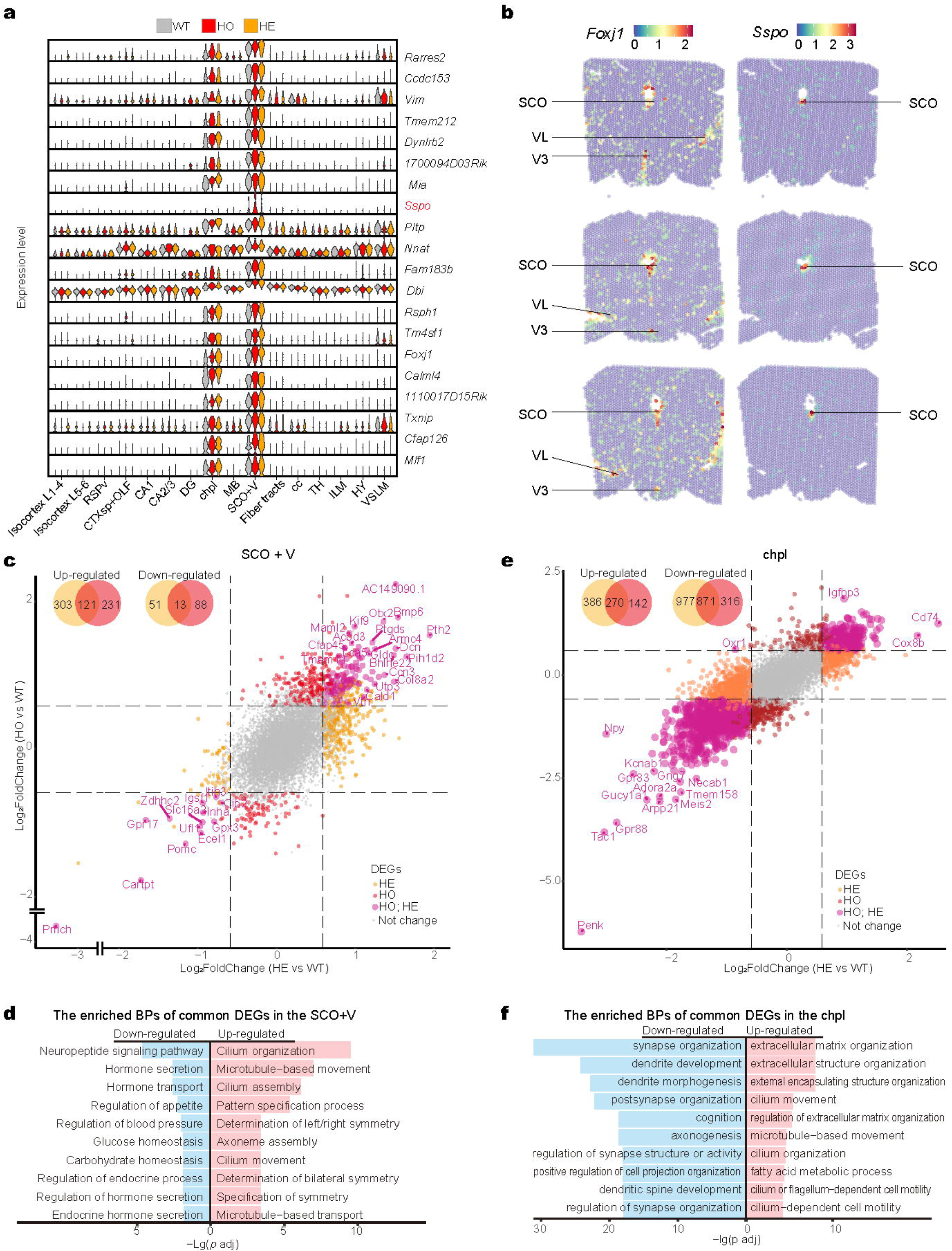
Differentially expressed genes in the SCO and ventricles are involved in CSF secretion, flow and homeostasis (See also Fig. S5) a. Top 20 genes with spatial specific expression in the spatial region “SCO + V”. The spots within the region “SCO + V” were compared with all spots in other regions to obtain the genes with spatial specific expression. b. The spatial expression of *Foxj1* and *Sspo*. *Foxj1* was specifically expressed in the SCO and the ventricles, and *Sspo* was exclusively expressed in the SCO. The scaled colors represent the normalized expression values in each spot. c. The DEGs of the spatial cluster “SCO + V”. The differential expression was calculated by comparing the expression values of the spots in the cluster “SCO + V” between the mutant mice and the wild-type mice. The two Venn diagrams in the up-left conner of the dot plots show the overlapping DEGs (up-, down-regulated respectively) between the heterozygous and homozygous mutants. Red, orange and magenta represent the DEGs of the homozygous (HO), the DEGs of heterozygous (HE) and the common DEGs, respectively. The common DEGs are shown as dots with larger sizes. The Y-axis shows log2(Fold Change) between the homozygous (HO) and the wild-type (WT) and X-axis show log2(Fold Change) between the heterozygous (HE) and the wild-type (WT). d. The enriched biological processes (BPs) of the common DEGs in the spatial cluster “SCO + V” of the heterozygous (HE) and homozygous (HO) mutant mice: left for the down-regulated DEGs and right for the up-regulated DEGs. The X-axis shows –lg (adjust *p* value). e. The DEGs of the spatial cluster “choroid plexus (chpl)”. See the legend for (c) for brief methods and explanations. f. The enriched biological processes (BPs) of the common DEGs in the spatial cluster “chpl” of the heterozygous (HE) and homozygous (HO) mutant mice: left for the down-regulated DEGs and right for the up-regulated DEGs. The X-axis shows –lg (adjust *p* value).

We looked into the gene expression change of the spots in the cluster “SCO + the ventricles (SCO + V)” of the mutant mice compared with the wild type mice: 488 DEGs (424 up-regulated and 64 down-regulated) in the heterozygous and 453 DEGs (352 up-regulated and 101 down-regulated) in the homozygous. The heterozygous and homozygous mutant mice share 121 up-regulated genes and 13 down-regulated genes in “SCO + the ventricles (SCO + V)” (**Fig. 6c**, **Table S3**). The functional enrichment analysis on the common DEGs revealed distinct functions of the up-regulated and down-regulated genes. The down-regulated genes are enriched in neuropeptide hormone secretion and transport functions (**Fig. 6d**, **Table S3**). Some of these down-regulated genes, such as *Pmch*, *Gpr50*, *Gpr17*, *Cartpt* (CART) and *Pomc*, are known to be specifically expressed in hypothalamus neurons, suggesting that the SCO and ventricles may have projection of neurons in the hypothalamus. The up-regulated genes are enriched in functions such as microtubule-based transport, movement, axoneme assembly, cilium organization, cilium assembly, cilium movement, etc. (**Fig. 6d**, **Table S3**), suggesting that there might be increased secretion activity and cilium movement of the SCO- and ventricular ependymal cells to recompensate the deficit of SCO-spondin. We looked into the known functions of some top up-regulated genes. BMP6 is an endogenous regulator of CSF hepcidin, an important player in brain iron homeostasis ^65^. PTH2 or parathyroid hormone related protein (PTHRP), a normal constituent of human CSF, is involved in intracellular calcium (Ca) regulation ^66^. These results suggest that SCO-spondin may play a crucial role in CSF secretion, flow and homeostasis.

We further looked into the gene expression change of the spots in the cluster “choroid plexus (chpl)”: 2,504 DEGs (656 up-regulated and 1,848 down-regulated) in the heterozygous and 1,599 DEGs (412 up-regulated and 1,187 down-regulated) in the homozygous. The heterozygous and homozygous mutant mice share 270 up-regulated genes and 871 down-regulated genes in “choroid plexus (chpl)” (**Fig. 6e**, **Table S3**). The down-regulated genes are mainly involved in neuronal development such as axonogenesis, dendrite morphogenesis, synapse organization, etc. (**Fig. 6f**, **Table S3**), suggesting that choroid plexus may be involved in neuronal development and that such function can be affected in the mutant mice. The up-regulated genes of the cluster in “choroid plexus (chpl)” have similar functional enrichment as those of the cluster “SCO + ventricles”: cilium organization and movement (**Fig. 6f**, **Table S3**), suggesting that there might be increased secretion activity and cilium movement of choroid plexus (chpl). In summary, the genes involved in regulating CSF secretion, movement and homeostasis, as well as neuronal morphogenesis and synapse organization, are dysregulated in the SCO, ventricles, and choroid plexus of the *SSPO*-PTV mutant mice, suggesting that SCO-spondin may serve as a signaling molecule mediating CSF-related functions through brain barriers.

### The genes involved in affective disorders are dysregulated in brain barrier cells of the mutant mice

To understand the cellular functional changes of the mutant brain at the single-cell level, we conducted single cell RNA-seq (SC-seq) on the whole brain (with cerebellum and medulla removed) of mice under normal condition, two mice for each genotype, using droplet-based 10× Genomics for library construction followed by Illumina sequencing (Methods). After removing doublets and low-quality cells that had extremely high/low UMI counts, gene numbers or mitochondrial ratios (Methods), we got 29,877 cells for the wild-type, 30,037 cells for the homozygous and 30,940 cells for the heterozygous **(Fig. S6a**). There are no shifts in gene expression patterns in the mutant mice compared with the wild-type mice (**Fig. S6b**, **Table S4**). Since different cell types have different UMI counts, gene numbers and mitochondrial gene ratios, we set different criteria of quality control for cleaning the data of different cell types (**Table S4**).

We used Seurat to integrate the SC-seq datasets of all 6 mice and annotated them using established cell-type markers for the brain cells ^67,68^. Based on the expression levels of the conserved markers of each cell type (**Fig. S6c**), 13 cell types were identified: endothelial cell (EC), microglial cell (MC), two subtypes of astrocyte (AC-1 and AC-2), two subtypes of oligodendrocyte (OL-1 and OL-2), neuron, ependymal cell (EpenC), choroid plexus epithelial cell (CPEpiC), olfactory ensheathing cell (OEC), smooth muscle cell (SMC), vascular leptomeningeal cell (VLMC) and macrophage (**Fig. 7a**). The UMI counts, gene numbers and mitochondrial gene ratios varied among different cell types, and didn’t show significant differences among the three genotypes (**Fig. S6d**).

**Fig. 7.**
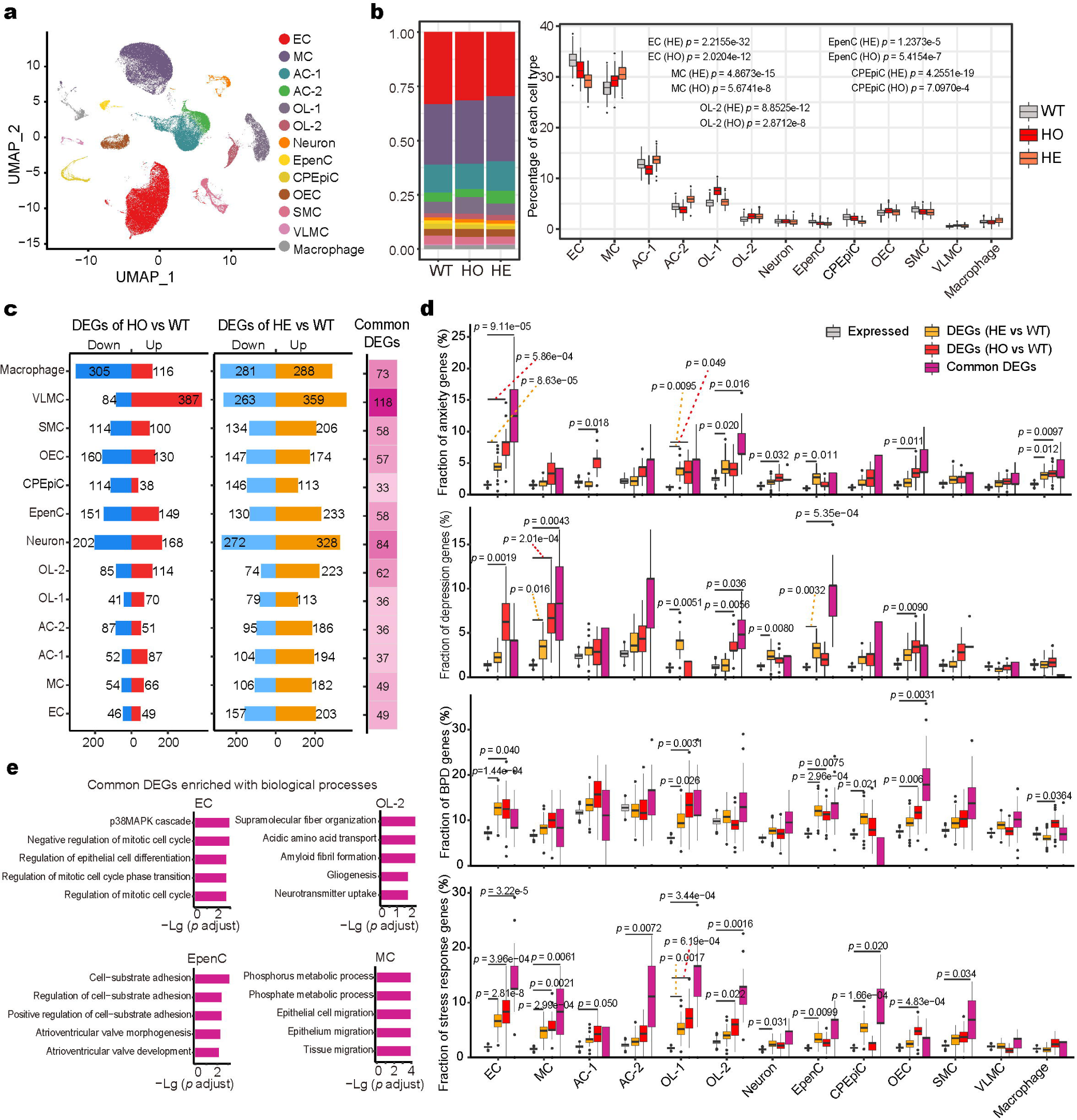
The differentially expressed genes in endothelial cells, ependymal cells, oligodendrocytes and microglial cells are enriched with genes involved in anxiety, depression and bipolar disorder (See also Fig. S6) a. UMAP embedding with the integration of the datasets of the three different genotypes (WT, HO and HE) using Seurat, colored by cell types. b. The percentage of cells per cell type of the three genotypes. The Y-axis shows the percentage of cells, calculated by random sampling 2% cells from the three genotypes respectively and the procedure was repeated 100 times. c. The DEGs of each cell type. The bar plot shows the number of DEGs. The red and blue bars show the up-regulated and down-regulated genes, respectively, in the homozygous (HO) mutant mice compared to the wild-type (WT) littermates. The light-blue and orange bars show the up-regulated and down-regulated genes of the heterozygous (HE) mutant mice compared to the WT littermates. The common DEGs of each cell type between the homozygous (HO) and the heterozygous (HE) are listed on the right. d. The gene set enrichment results for the DEGs of each cell type. The enrichment analysis was performed on the candidate gene sets for anxiety, depression, bipolar disorder and stress response for the cell-type DEGs of the heterozygous (HE), the homozygous (HO) and the common DEGs. The background genes are the expressed genes in each cell type. e. The enriched biological processes (BPs) of the common DEGs in four cell types: ECs, OL-2, EpenCs and MCs. Only the top five enriched BPs are shown. The bar shows −lg (adjusted *p* values).

We compared the normalized cell numbers of these cell types between the mutant and the wild-type mice, and found that ECs, CPEpiCs and EpenCs decreased in numbers in the mutant mice, while MCs and OLs increased in numbers (significant increase in the homozygous and an increasing trend in the heterozygous) (**Fig. 7b**). EpenCs are known to participate in the production, regulation and circulation of CSF and associate with anxiety and depression ^69^. ECs constitute the CSF-blood barriers and CSF-brain barriers and are known to associate with mood disorders ^70^. The CPEpiCs provide secretory functions and maintain ionic gradients across the blood-CSF barrier and mediate physiological immune-brain communication ^71^ and also associate with mood disorders ^72^. Interestingly, both microglial proliferation and oligodendrocyte proliferation are known to participate in stress response ^73,74^, probably via regulating the permeability of the blood-brain barriers ^75,76^.

To evaluate the gene-expression change in each cell type in the mutant mice, we calculated DEGs for each cluster using Seurat, and defined the DEGs under the cutoff *p* < 0.05 and |fold change| > 1.1 (**Fig. 7c**, **Table S4**). We also mapped the DEGs obtained from bulk RNA-seq on the whole brain to SC-seq data, and found that each cell type had a different contribution to the bulk DEGs. ECs, EpenCs, MCs and macrophages contributed more to the bulk DEGs of the mutant mice than other cell types (**Fig. S6e**). For each cell type, we checked the enrichment of the DEG sets (of the heterozygous, the homozygous and the common DEGs) in the candidate gene sets of affective disorders (anxiety, depression and bipolar disorder) and stress response, respectively. The ECs showed enrichment with the candidate gene sets of anxiety, bipolar disorder and stress response in both the homozygous and heterozygous mutant mice, and enrichment with the depression candidate genes in the homozygous (**Fig. 7d**). At least one of OL populations (OL-1, OL-2) showed enrichment with candidate gene sets of stress response genes, anxiety, depression and bipolar disorder in the homozygous and heterozygous mutant mice, respectively (**Fig. 7d**). The EpenCs showed enrichment with stress response genes for both mutant genotypes, and enrichment with the candidate gene sets of anxiety disorder and depression for the heterozygous mice (**Fig. 7d**). MCs showed enrichment with depression candidate genes and stress response genes for both genotypes (**Fig. 7d**). The OECs showed enrichment with the candidate gene sets of the anxiety, depression and stress response for the homozygous mice (**Fig. 7d**), while CPEpiCs showed enrichment with the candidate genes of bipolar disorder and stress response for the heterozygous mice (**Fig. 7d**). The enrichment of candidate gene sets of affective disorders suggest that these cell types may be involved in the affective symptoms of the schizoaffective disorder.

We further checked functional enrichment of the DEGs of each cell type to explore the affected biological processes (**Fig. 7e** and **Fig. S6f**). We were particularly interested in the affected functions of ECs, EpenCs, CPEpiCs, OLs and MCs. ECs are mainly enriched with cell cycle regulation functions; EpenCs, cell−substrate adhesion regulation functions; CPEpiCs, ion homeostasis; OLs, transmitter transportation functions; MCs, metabolic processes and cell migration (**Fig. 7e**). These results are consistent with previous studies on the roles of these cell types in affective disorders. EC is the major cell type of CSF-blood barrier and CSF-brain barriers, and the differentiation of ECs is associated with depression ^77^; cell adhesion is critical for EpenC to constitute CSF-brain barrier, which is associated with depression ^69^; the ion homeostasis in the CPEpiCs maintains ionic gradients across the blood-CSF barrier ^71^ and is involved in depression ^69^; OLs play an important role in regulating the endothelial cells of blood-brain barriers ^76^, the transmitter transportation function of OLs is involved in mood disorders ^78^; the metabolic status of MCs supports immune surveillance of the brain ^79^, which may play a role in mood disorders ^80^. These results suggest that the disrupted pathways in the cell types involved in the brain barrier-related functions may underly affective symptoms in the schizoaffective disorder.

### The cell-cell communications such as WNT signaling involving brain barriers are dysregulated in the mutant mice

We speculated that after being secreted into CSF, SCO-spondin or its derived peptides may regulate cellular functions by interacting with proteins. To find the interactors of SCO-spondin, we searched for the proteins interacting with SCO-spondin, using physical interaction data from the STRING database ^81^ and protein complex data from CORUM ^82^. The direct interactors of SCO-spondin include the proteins PTK7, NTRK3, LRRK2, DAG1, ROR1, MET, MST1R, NTRK1, ROR2, LRRK1, UTRN, DMD (**Fig. S7a**, **Table S5**), which are considered as potential receptors. We checked the expression of these genes (**Fig. S7b**) and found that *Utrn*, *Dag1*, *Dmd* and *Ptk7* were expressed in cell types including EpenCs, ECs, CPEpiCs or VLMCs (**Fig. S7b**), which constitute brain barriers. The proteins encoded by these four genes are cell surface proteins, indicating that these SCO-spondin interactors on the brain barrier cells may transduce the signal.

We further analyzed cell-cell communication pathways between these 13 cell types using CellChat (**Fig. 8a**, Methods). Interestingly, there were more altered pathways originated from the cell types constituting the blood-brain barriers (i.e. VLMCs, CPEpiCs, SMCs and ECs) and the CSF-brain barrier (i.e. EpenCs) (**Fig. 8a**, **Table S5**). More signaling pathways targeting OLs, OECs, VLMCs, ECs and EpenCs were altered in both mutant mice (**Fig. 8a**, **Table S5**): OLs received increased signaling from the VLMCs, ECs, EpenCs, SMCs and OECs in both mutant mice, while the OECs receive decreased signaling from these cell types (**Fig. 8a**). The ACs also received decreased signaling from these cell types in the heterozygous (**Fig. 8a**). We also measured the relative strength of the signaling transduction in the wild-type and mutant mice, and thus deduced the turn-on/off or the increase/decrease in the mutant mice compared to the wild-type mice according to the activity status of the ligand-receptor pairs (**Fig. S7c**). The total received signaling strengths in ECs, MCs, ACs, OLs and OECs were altered more than other cell types (**Fig. S7c**, **Table S5**), while the total source signaling strengths in ECs, MCs and ACs were altered more than others (**Fig. S7c**, **Table S5**). These results support the conclusion that the cell-cell communications involving brain barrier cells are affected by the deficit of SCO-spondin in CSF of the mutant mice.

**Fig. 8.**
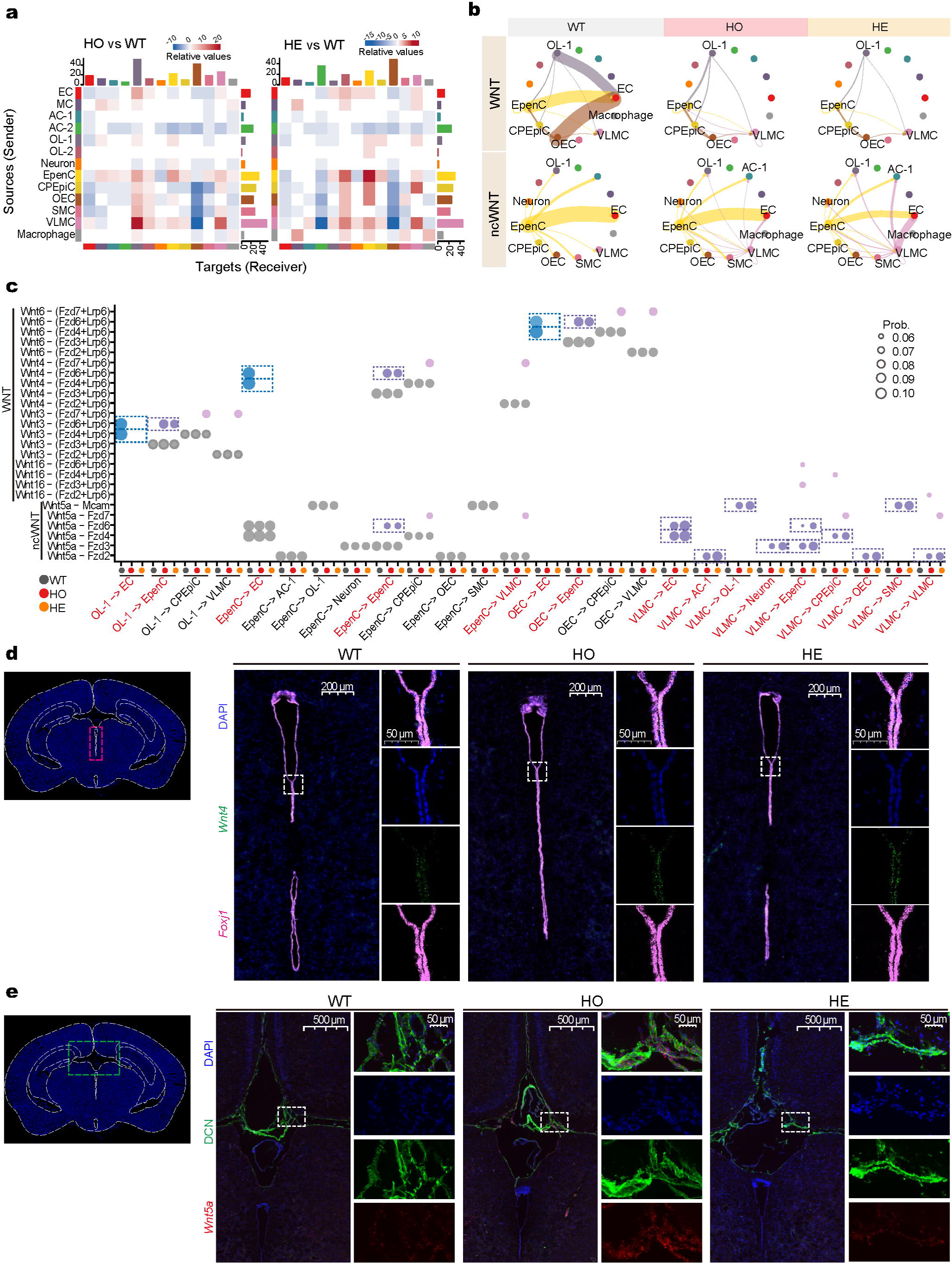
Abnormal cell-cell communications in the mutant mice (See also Fig. S7). a. The number of differential interactions in the cell-cell communication networks between the mutant and wild-type datasets. The colored bars on top represent the sum of incoming signaling values of the column in the heatmap. The colored bars on right represent the sum of row outgoing signaling values. The scaled red/blue colors in the heatmaps represent the increased/decreased signaling between the cell types of the mutant mice compared to the wild-type mice. b. Canonical and non-canonical WNT pathways for the cell-cell communications in the wild-type (WT), homozygous (HO) and heterozygous (HE) mice. Edge colors are consistent with the signaling sources, and edge weights are proportional to the signaling strengths. c. The activated ligand-receptor pairs in canonical WNT and non-canonical WNT signaling pathways. The dot size shows the communication probability. The X-axis shows the signals sent from source cells to target cells. Grey, orange and red show source-to-target cell-type pairs in the wild-type (WT), the homozygous (HO) and the heterozygous (HE), respectively. Red font shows the cell-type pairs with signals turned on/off in the mutant mice. The red dotted boxes highlight the consistent changes of the ligand-receptor pairs in both the homozygous (HO) and the heterozygous (HE). d. The expression of *Wnt4* was detected in the ependymal cells of the wild-type (WT), homozygous (HO) and heterozygous (HE) mice using RNAscope. *Wnt4* mRNAs were probed with RNAscope probes (Green). *Foxj1* mRNAs were probed with RNAscope probes (pink) to label the ependymal cells. The nuclei were stained with DAPI (blue) for recognition of cell body positions. e. The expression of *Wnt5a* was detected in VLMCs in the wild-type (WT), homozygous (HO) and heterozygous (HE) mice using RNAscope. *Wnt5a* mRNAs were probed with RNAscope probes (Red). DCN proteins were probed with anti-DCN antibody (Green) to label the VLMCs. The nuclei were stained with DAPI (blue) for recognition of cell body positions.

We identified 10 signaling pathways with altered signaling strengths (**Fig. S7d**), and visualized the changes of these 10 signaling pathways with significantly altered strengths between the 13 cell types (**Fig. 8b**, **Fig. S7e**). Four pathways (WNT signaling in ECs; ncWNT signaling in VMLCs; MIF signaling in ECs, EpenCs and SMCs; CXCL signaling in EpenCs) have similar change patterns between the homozygous and the heterozygous (**Fig. 8b** and **Fig. S7e**). The WNT and ncWNT signaling are decreased between EpenCs and ECs in the mutant mice, while ncWNT signaling is increased between VLMCs and ACs, OLs, neurons or other cell types (**Fig. 8b**). CXCL signaling is decreased between ECs and EpenCs or SMCs (**Fig. S7e**). The MIF signaling is increased between EpenCs, ECs and MCs, and between SMCs and OLs, MCs, ECs, OECs or CPEpiCs (**Fig. S7e**). These four signaling pathways are documented to associate with stress response, anxiety and depression ^83–86^. These findings on cell-cell communications suggest that EpenCs, ECs, MCs and OLs may contribute to the mood-related phenotypes of the *Sspo* PTV mutant mice through WNT, ncWNT, MIF and CXCL signaling pathways.

We checked the expression of WNTs and their receptors in the 13 cell types, and found 21 WNT-to-receptor pairs that had similar change patterns in both the heterozygous and the homozygous (**Fig. 8c**). WNT4 mediates signaling originating from EpenC, WNT5a from EpenC and VLMC, WNT3 from OL-1 and WNT6 from OEC (**Fig. 8c**). WNT3, WNT4 and WNT6 signaling from EpenC, OL and OEC to EpenC were turned on, while these signaling from EpenC, OL and OEC to EC are turned off. The WNT5a signaling from VLMC to several other cell types was turned on (**Fig. 8c**). We carried out RNAscope to verify the expression of *Wnt4* in EpenCs (**Fig. 8d**) and the expression of *Wnt5a* in EpenC and VLMC (**Fig. 8e**). WNT signaling pathways are reported to be essential in the development, maintenance and repair of blood-brain barriers ^87–89^. These multiple dysregulated WNTs-mediated cell-cell communications involving brain barrier cells strongly support SCO-spondin as a regulator of brain barriers, and the deficit of SCO-spondin may cause brain barrier dysfunction leading to abnormal mood-related behaviors.

## Discussion

Psychiatric disorders, such as schizophrenia and affective disorders, are complex diseases and are highly polygenic, with each risk variant conferring different sizes of effects ^90^. The high heterogeneity of genetic factors and the interactions between the genetic factors generate spectrums of phenotypes, resulting in an enormous challenge in their classification ^91^. The genetic studies on families strongly suggest that schizoaffective disorder as interform between independent genetic factors of schizophrenia and an affective disorder ^5^. We identified 4 potential causal genes (*SSPO*, *FGFRL1*, *ASPN and OR13C5*) with protein truncating variants (PTVs), fitting Mendelian dominant inheritance model, in the schizoaffective family (**Fig. 2a, S1a**, **Table S1**). *SSPO* is reported in CNVs of patients with bipolar disorder ^35^ and is associated with irritable bowel syndrome and depressive disorder ^37^ and schizophrenia ^38^. The mouse model of *SSPO* PTV demonstrated mood-related phenotypes, but no psychotic-related phenotypes (**Fig. 3**), suggesting that the *SSPO* PTV may contribute to affective symptoms while other genes may be responsible for schizophrenic symptoms. Interestingly, of the other three genes with PTVs, *FGFRL1* and *OR13C5* are associated with schizophrenia ^43,44^. Considering the high polygenicity of schizophrenia and affective disorders, such co-occurrence of independent genetic factors may not be an uncommon phenomenon.

SCO-spondin, encoded by *SSPO*, is exclusively expressed in the SCO (**Fig. 2d** and **Fig. 6a-b**). The SCO is the first structure to differentiate in embryo development and remains fully developed and active during whole life in most vertebrates ^92^. In human, it is morphologically distinguishable in 7-to 8-week-old embryos, shows active secretion in 3-to 5-month-old fetuses, and regresses to a few islets of secretory ependymal cells in 9-year-old children ^92^. Although the functional importance of the SCO has been intriguing to researchers for more than half a century ^13^, its function remains unknown. The mood-related phenotypes of *Sspo* PTV mice (**Fig. 3** and **Fig. S2**) suggest a function in mood regulation for this mysterious organ. The SCO belongs to the circumventricular organs (CVOs), which are located around the third and fourth ventricles, mediating the communication between the brain and the periphery by performing sensory and secretory roles, with some of them (such as the pineal organ and the area postrema) known to regulate mood, sleep and food intake ^93^.

Multilayers of transcriptomics consistently reveal that the differentially expressed genes (DEGs) are relevant to the affective disorder-like behaviors in the brain of the *Sspo* PTV mutant mice (**Fig. 4g-h**; **Fig. 5d**; **Fig. 7d**). The DEGs of the whole brain, identified by bulk transcriptomics, are enriched with candidate gene sets of stress response, anxiety, depression and bipolar disorders (**Fig. 4g-h**), but not with candidate gene sets of SCZ and ASD (**Fig. S3D-E**), suggesting that the *SSPO* PTV may play a role in mood disorders. The DEGs of CSF-contacting brain regions, revealed by spatial transcriptomics, are enriched with the same gene sets (**Fig. 5c-d**). Of these CSF-contacting regions, the SCO, ventricles and choroid plexus have direct contact with CSF through ependymal cells, while olfactory area and hypothalamus are known to have CSF-contacting neurons ^60,61^, suggesting that the deficit of SCO-spondin caused by the PTV mutation may be involved in the abnormal mood-related behaviors. Furthermore, the DEGs of cell types EpenCs, ECs, OLs and MCs are enriched with the above candidate gene sets, respectively (**Fig. 7c-d**), suggesting that these cell types may be involved. EpenCs form the CSF-brain barriers of the ventricles and constitute the CSF-blood barriers of choroid plexus, and ECs constitute the CSF-blood barriers of choroid plexus in the ventricles and blood-brain barriers between blood and brain parenchyma, while OLs and MCs have direct contact with the brain barrier cells and regulate the brain barrier integrity ^75,76,94,95^. Abnormal behaviors and affective disorders have been reported to be associated with the disruption of the brain barriers ^96,97^.

The dysregulated functions of the DEGs, revealed by the enriched GO biological processes of the DEGs, converge at functions relevant to brain barriers, such as barrier development, formation, transportation, integrity, CSF secretion and CSF movement. The DEGs of the whole brain are enriched with functions of epithelium development, stress response and immune regulation (**Fig. 4i-j**). The DEGs of the CSF-brain barriers and CSF-blood barriers are enriched with functions of cilium organization, cilium movement and secretion (**Fig. 6d-f**). It’s worth mentioning that the neuronal development function is also affected in the CSF-blood barriers (**Fig. 6f**). The DEGs of the EpenCs are enriched for functions of CSF-brain barrier formation and integrity such as cell adhesion (**Fig. 7e**), while the DEGs of ECs are enriched functions for CSF-blood barrier formation and integrity such as epithelial proliferation and differentiation (**Fig. 7e**). In addition, the DEGs of OLs are enriched for functions of extracellular supramolecular organization and acidic amino acid transportation (**Fig. 7e**) and the DEGs of MCs are enriched with epithelial cell migration (**Fig. 7e**). Therefore, the affected brain regions and cell types converge at CSF-brain barriers and CSF-blood barriers and two other brain regions which have neurons contacting CSF, suggesting that the SSPO PTV may contribute to the affective symptoms of schizoaffective disorder through CSF and brain barriers.

These affected barrier cells, in turn, change the components and homeostasis of the CSF and the cell-cell communication between themselves and other cell types. The four most affected cell-cell communication signaling pathways are WNT, ncWNT, MIF and CXCL signaling pathways involving barrier cells (**Fig. 8** and **Fig. S7**). WNTs mediate cell-cell communications from EpenCs, OLs and OECs to ECs and VLMCs to several cell types, and CXCL mediates communications from ECs and VLMCs to EpenCs and SMCs, while the MIF mediates communications from ECs, CPEpiCs, OLs, MCs and OECs to SMCs and EpenCs. WNT signaling pathways are reported to be essential in the development, maintenance and repair of blood-brain barriers ^87–89^. Chemokines CXCLs can disturb brain barrier morphology and function ^98^. The cytokine MIF enhances blood-brain barrier permeability ^99^. Disruption of the brain barriers is reported to be involved in affective disorders ^96,97,100^.

Overall, our results on the *SSPO*-PTV animal model (**Fig. 2**) support the PTV mutation as a genetic factor for the affective symptoms of schizoaffective disorder (Fig. 3), and reveal affected brain regions and cell types through multilayers of transcriptomics. Spatial transcriptomics identified the SCO, brain barriers, cortical subplate and olfactory cortex and hypothalamus, which have cells or neurons contacting CSF, as the key affected brain regions involved in the mood-related behavioral phenotypes of the mutant mice (**Fig. 5-6**). Single cell RNA-seq reveals the brain barrier cells as the most affected cells (**Fig. 7-8**). The identification of CSF-contacting brain barriers and brain regions as the most relevant regions indicates that the CSF should be an important mediator in the development of the mood-related behaviors of the mutant mice. It is reasonable that these CSF-contacting brain regions are the most affected regions because the SCO secretes SCO-spondin into the CSF. The deficit of the SCO-spondin in the CSF can directly regulate accessible cells through binding to SCO-spondin receptors and induce functional changes in the brain barrier cells. This study reveals, for the first time, the function of the SCO in stress response and mood regulation through secreting SCO-spondin into CSF, and the link of its deficit due to a PTV with affective disorders.

This work answers the important question about whether genetic factors contribute, independently, to the affective symptoms and schizophrenic symptoms of schizoaffective disorder, and demonstrated that a PTV in *Sspo* resulted in mood-related behaviors in the mouse model. The protein SCO-spondin may affect the brain through CSF. Given its high conservation and its potential interactions with cell surface receptors, SCO-spondin may regulate cells contacting CSF. The CSF-contacting brain regions are significantly affected, and the dysregulated genes are involved in stress response and mood disorders. Thus, what precisely underlies the initial effects of SCO-spondin deficit on the brain barriers and whether it can be a therapeutic target for the affective symptoms should be an active area of investigation. Considering the exclusive expression of SCO-spondin in the SCO, understanding the function of SCO-spondin would also retrigger the curiosity about the function of this mysterious organ in the brain.

### Limitations of study

We have identified multiple genes with PTVs in the patients of the schizoaffective family, but we have only generated a mouse model for the PTV in *SSPO*. The interactions between the mutations of different genes might contribute to those of the symptoms found in the patients of the family. More studies are urgently needed to illustrate the genetic interactions in the complex disease.

## Data Availability

All data produced in the present study are available upon reasonable request to the authors

## Acknowledgements

The work was supported by Regional Joint Fund of Guangdong Province (Grant No. 2019B1515120080), the Key Scientific and Technological Projects of Guangdong Province (Grant No. 2018B030335001), the National Natural Science Foundation of China (Grant No. 81571097), and the Natural Science Foundation of Guangdong Province (Grant No. 2016A030308020). The funding organizations had no role in design and conduct of the study; collection, management, analysis, and interpretation of the data; or preparation, review, or approval of the manuscript.

## Author contributions

X.Y. conceived the project; Y.G. carried out the computational analysis; Y.G., J.X, S.L., L.X. and W.Y. acquired the experimental data. F.F and B.Z performed the diagnosis of the family with schizoaffective disorder. Y.G. and X.Y. wrote the manuscript. All of the authors contributed to the preparation of the manuscript.

## Methods

### DNA preparation

Peripheral blood samples were collected with EDTA-Na2 anticoagulant blood collection tubes. The genomic DNA was extracted from the blood using Blood DNA Mini Kit (Magen Biotech, Guangzhou, China) according to the manufacturer’s protocol. DNAs was assessed using a NanoDrop spectrophotometer (Thermo Fisher Scientific, Waltham, MA, USA).

### Exome sequencing and data analysis

Whole exome sequencing was carried out on the Illumina HiSeq X Ten NGS platform with libraries prepared using Illumina exome kits by Berry Genomics (Beijing, China). Reads were aligned with the reference human genome (Homo_sapiens.GRCh38.85) using the Burrows-Wheeler algorithm tool (BWA, version 0.7.15). The picard pipeline (version 2.2.2) was used to perform additional sequence QC checks and compute quality metrics to create final BAM files. Genome Analysis Toolkit (GATK, version 3.5, best practices) was used to call single-nucleotide variants (SNVs) and small insertion/deletion (INDEL) variants from individual BAM files, and recalibrated variant quality scores (VQSR) were generated to determine the overall variant quality in the VCF (variant calling format) file. ANNOVAR (version 2016-02-01) (with the configuration: --protocol refGene, cytoBand, esp6500siv2_all, 1000g2015aug_all, avsnp150, dbnsfp30a) was used to annotate the merged VCF files. The potential pathogenic SNVs and indels were selected using the following criteria: (1) variants that might cause amino acid alterations in the proteins; (2) variants are found in all the patients, but not in normal individuals in the family.

### Generation of *Sspo* PTV knock-in mice

The C57BL/6 knock-in (KI) mouse model for the PTV mutation in *Sspo* gene was created by CRISPR/Cas-mediated genome engineering technique. The mouse *Sspo* gene (GenBank accession number: NM_173428.3; Ensembl: ENSMUSG00000029797) has 103 exons, with the ATG start codon in exon 1 and TGA stop codon in exon 103. The mutation target site is in Exon 76. A gRNA targeting vector with a donor oligo (targeting sequence flanked by 120 bp homologous sequences on both sides) was designed. An additional sequence (one base: A) in donor oligo was introduced into exon 76 by homology-directed repair. A silent mutation (AGC>AGT) was introduced to prevent the binding and re-cutting of the sequence by gRNA after homology-directed repair. Cas9 mRNA, gRNA generated by in vitro transcription and donor oligo were co-injected into fertilized eggs for KI mouse production. The pups were genotyped by PCR followed by Sanger sequencing.

### Experimental animals

The mice were housed 2–5 per cage under standard conditions (12 h/12 h light/dark cycle, access to dry food and water ad libitum). All of the experiments were conducted in accordance with the Regulations for the Administration of Affairs Concerning Experimental Animals (China) and were approved by the Southern Medical University Animal Ethics Committee.

The experiments were performed on 8-to-12-week male mice. The genotypes of heterozygote, homozygote and wild-type were determined by PCR amplification of the target sequence followed by Sanger sequencing. Age-matched male littermates were randomly assigned to each experiment. All the experiments were performed and analyzed blind to genotyping or group allocation.

### Immunofluorescent assay

SCO-spondin polyclonal antibodies were raised against the peptides deduced from the mouse SCO-spondin by injecting rabbits with antigen peptides derived from mouse the protein sequences (Sino Biological Inc., Beijing, China). The two peptides (HVEEEVTPRQEDLVPC, CPGPGIWSSWGPWEK) are designed, one according to the sequence close to the N-terminal of the SCO-spondin protein, and another just before the PTV site.

Mice were anesthetized by intraperitoneal injection (IPI) of 4% chloral hydrate/0.75% sodium pentobarbital (0.1ml/10g). During the mouse perfusion, saline will pass into the mouse circulation for 2-5 minutes. Subsequentially, the brain was immediately extracted, placed in ice-cooled PBS and washed twice with PBS. The whole brains were embedded in OCT (Optimum cutting temperature compound) cryostat sectioning medium and stored at -80°C. The brain was cut into 10-15 um thick slices by vibratome (LEICA CM1950), which were mounted onto glass slides. Slides were warmed to room temperature, fixed in 4% paraformaldehyde for 30-45 minutes, washed 3 times with PBS for 2 minutes each, and then incubated with 0.3% Triton-x-100 for 20 minutes and washed 3 times with PBS 2 minutes each. A circle was drawn on the slide around the tissue slice with a hydrophobic barrier pen. The tissue slices were incubated with 5% BSA (bovine serum albumin) for 30 minutes at room temperature to blocked non-specific binding. SCO-spondin polyclonal antibodies were diluted in PBS (1:100) and added onto the tissue slices, incubated and stored overnight at 4°C. The next day, the tissue slices were washed 3 times with PBST for 5 minutes. A fluorescent labeled secondary antibody (goat anti-rabbit IgG H&L, Alexa Fluor 488) diluted in 1:2500 PBS was incubated with the slices at room temperature for 1 hour. Slices were washed 5 times with PBST for 2 minutes each. After DAPI staining, a coverslip was placed on top of the tissue slices, and the edges of the coverslip was sealed with clear fingernail polish to prevent the cells from drying. Slides were examined under a fluorescence microscope (Imager D2) with the appropriate fluorescent filter sets.

### Behavioral test procedures

All behavioral test procedures were performed on adult male mice. On the test day, mice were transferred to the testing room and acclimated to the room conditions for at least 1 hour. After each individual test session, the apparatus was thoroughly cleaned with 75% alcohol to eliminate the odors and traces of the previous tested mouse. The tests were performed by experimenters who were blinded to the experimental group between 1:00 and 6:00 p.m. All data were shown as mean ± SEM and analyzed using unpaired t test, one-way ANOVA or two-way ANOVA. All the statistical analyses were done using Prism 7 (Graph Pad). Open field test, marble-burying, elevated plus maze, pre-pulse inhibition test, Y-maze, tail suspension test and learned helplessness test are described below.

### Open field test

Each individual mouse was placed facing the wall of 40 cm L × 40 cm W open field arena, and the movement of the mouse was recorded by Versamax Animal Activity Monitor (AccuScan Instruments, Inc, Made in USA) for 30 minutes as previously described ^101^. The recorded video file was further analyzed by video tracking software (Versamax Dat Version). Total distance traveled and time in the center area were measured. The open field was cleaned with 75% ethanol and wiped with paper towels between each trial.

### Elevated plus maze

The test consists of an elevated, plus-sign-shaped runway that was approximately 40 cm above the floor, with two opposing wall-closed arms (50 cm L × 10 cm W) and two opposing open arms (50 cm L × 10 cm W), and an intersection (10 cm L × 10 cm W) between the arms. At the time of the test, each mouse was placed in the center of the intersection, facing the closed arm. Each test was videotaped for 5 minutes. The time spent in the closed and open arms was quantified autonomously by the computational software.

### Marble burying

Mice are placed for thirty minutes in a standard cage filled with 5 cm depth of wood chip bedding in 5 rows of 4 marbles evenly spaced. After 30 minutes, the buried marbles were counted. A marble was considered buried if two-thirds of the marble is covered with bedding.

### Pre-pulse inhibition

The startle reflex measurement system (SR-LAB, USA) was used to measure startle response to a loud noise and pre-pulse inhibition of the startle response. A 20-minutes test session was initiated by placing a mouse in a plastic cylinder in a sound-attenuating chamber, in which the mouse was left undisturbed for the first 10-minutes period. The mouse was then subjected to startle-stimulus-only trials and pre-pulse-pulse trials for 10 minutes. White noise (40 ms) was used as the startle stimulus for all trial types. The startle response was recorded for 400 ms starting with the onset of the startle stimulus. The background noise level was 70 dB. The peak startle amplitude was used as a dependent variable. The intensity of the startle stimulus was 120 dB. A pre-pulse sound, with intensity of 74, 78, 82, 86 or 90 dB and duration of 20 ms, was randomly presented 100 ms before a startle stimulus. It also accepts no stimulations as one trial. Six blocks of the seven trial types were presented in a pseudorandom order such that each trial type was presented once within a block. The average inter-trial interval was 15 s (range: 10-20 s). All data shown are means ± SEM and analyzed using repeated unpaired t test.

### Y-Maze test

Mice were tested for spontaneous alterations using Y maze. In each trial, a mouse was placed in the center of the Y maze and left to roam freely until 8 minutes had elapsed. Three consecutive choices in three different arms were considered an alteration. The average percentage of alterations was scored.

### Tail suspension test

The tail suspension test (TST) was conducted using tail suspension cubicles (PHM-300, MED-Associates)^102^. Mice were individually suspended by the tail to a metal hanger with adhesive tape affixed 1.0-1.5 cm from the tip of the tail for 6 minutes. Data were collected using Med Associates Tail Suspension software. The time of the immobility period for the total 6-minutes TST session was summed from the recorded video for each mouse.

### Learned helplessness test

The learned helplessness procedure was conducted following the previously described procedures ^51^. Mice were placed on one side of a shuttle box that was divided into two equal chambers with an auto-controlled guillotine door. Inescapable foot shocks were delivered to mice repeatedly 100 times at an amplitude of 0.3 mA, for a duration of 5 s and a randomized inter-shock interval of 5-99 s over 3 consecutive days. Non-shocked mice were exposed to the chamber for the same period of time without receiving shocks. Twenty-four hours after the third shock procedure, learned helplessness was assessed by testing escape performance, which consisted of 30 escape trials. Each trial adopted a single 0.3 mA foot shock administered for a maximum duration of 24 s with randomly varying inter-trial interval (range = 30-60s). During the first five trials, the gate opened as the shock was administered. For the remaining 25 trials, the gate opened 2 s after the shock, which facilitates the observation of helplessness. Each trial was terminated when either the mouse escaped to the non-shock side of the shuttle box or the maximum duration (24 s) was reached. The mean escape latency and the total number of escape failures over the final 25 successive escape trials were presented.

### RNA preparation

For each group (wild-type, heterozygous and homozygous) without undergoing stress, six whole-brain RNA samples were prepared, and for each group (wild-type, heterozygous and homozygous) after the stress of the tail suspension test, five whole-brain RNA samples were prepared. Each RNA sample was extracted from the whole brain of one adult mouse according to the manufacturer’s protocol (RNAeasy Mini Kit, Qiagen, USA). The quality and yield of the isolated RNAs were assessed using a NanoDrop Spectrophotometer (Thermo Fisher Scientific, Waltham, MA, USA) and Agilent 2100 Bioanalyzer (Agilent Technologies, Santa Clara, CA, USA). Only RNA samples with a high RNA integrity number (RIN > 9) were selected and used for the subsequent sequencing.

### RNA-Seq data analysis

RNA sequencing was performed at Berry Genomics (Beijing, China) using Illumina NovaSeq. Sequenced reads were mapped to mouse whole genome (GRCm38) using STAR_2.5.2b. Raw counts were calculated by HTSeq-count (version 0.11.2). Pair-wised analyses were performed, and genes with > 3 raw counts in each group were considered as expressed genes and used for downstream analysis (**Table S2**). The principal components analysis (PCA) was performed by plotPCA function using normalized count by variance stabilizing transformation function of DEseq2 (v1.20.0). An outlier in wild-type group which underwent stress was excluded in the downstream analysis.

Differential expression analysis on two groups was performed using the DESeq2. Differentially expressed genes (DEGs) were determined using a cutoff of *p* < 0.01 and absolute fold change |FC|≥1.3 for DESeq2 (**Table S2**).

The threshold-free comparison of differential gene expression between different groups of mice was carried out using R package “RRHO2” (version 1.0). A Full threshold-free gene list of differential transcriptomes (HO mice or HE mice compared to WT mice) were ranked by the –lg (*p* value) multiplied by the sign (-or +) of the fold change from the DESeq2 analysis. The lg-transformed hypergeometric *p* value are plotted to map the degree of the statistically significant overlap between two differential transcriptomes (two ranked gene lists). Each pixel in a RRHO map represents the signed lg-transformed hypergeometric *p*-value for the overlap of subsections of two ranked lists from a point on the map (corresponding to a rank threshold pair: Rank X, Y) to either bottom left corner (Rank 1, 1) or top right corner (Rank N, N). Genes along each axis are sorted from the most significantly up-regulated to the most down-regulated from the lower left corner.

### Disease candidate genes used for enrichment analysis

#### 1. The candidate genes of anxiety

To search for candidate genes from literature, the keyword “anxiety” was used to retrieve papers from PubMed database (https://www.ncbi.nlm.nih.gov/pubmed/), and 205 genes from 31 publications were manually annotated.

#### 2. The candidate genes of depressive disorder

The keyword “depression” was used to retrieve Genetics Home Reference dat abase (http://ghr.nlm.nih.gov/), polygenicpathways database (http://www.polygen icpathways.co.uk/depression.htm), and PubMed database (https://www.ncbi.nlm. nih.gov/pubmed/), and 300 genes were collected.

#### 3. The immune candidate genes

All 3,904 immune genes used are from our previously collected immune gene (Gao et

al., *iScience*, 2021 Oct 2;24(11):103209. doi: 10.1016/j.isci.2021.103209).

#### 4. The candidate genes of bipolar disorder

The differentially expressed genes of bipolar patients were identified by (Akula N et al., *Mol Psychiatry,* 2014 Nov;19(11):1179-85. doi: 10.1038/mp.2013.170), and 2,066 DEGs were use in the enrichment analysis.

#### 5. Stress response candidate genes

All 364 Stress response genes used are from literature (Flati T et al., *Sci Data.* 2020 Dec 16;7(1):437. doi: 10.1038/s41597-020-00772-z, TableS1)

#### 6. The candidate genes of autism spectrum disorder

All 1,036 candidate genes of autism spectrum disorder (ASD) were collected from Autism database (http://autism.mindspec.org/autdb/HG_Home.do).

#### 7. The candidate genes of schizophrenia

All 1,592 candidate genes of schizophrenia used are from our previous publication (Gao et al., *iScience*, 2021, TableS3-2).

## Spatial transcriptome sequencing

### Sample preparation

The coronal slices of the frozen brain of an adult mouse of each genotype were prepared for spatial transcriptome sequencing. Both the tissue block and the Visium slide were equilibrated inside the cryostat at -20 °C for 30 minutes before cryosectioning. The coronal slices through the entire brain were cut at a thickness of 10 µm and immediately placed on the Visium array slide (Visium Spatial Gene Expression slides, 10× Genomics).

Samples were processed according to the Visium Spatial Gene Expression User Guide (10× Genomics) and all reagents were from the Visium Spatial Gene Expression Kit (10× Genomics). Briefly, sections were fixed in chilled methanol at −20 °C for 30 minutes, stained with hematoxylin and eosin, and mounted in 85% glycerol for imaging. Olympus TX83 microscope at 10× magnification was used for imaging. The slices were then permeabilized at 37 °C for 12 minutes. After permeabilization, the on-slide reverse transcription (RT) reaction was performed at 53°C for 45 minutes. Permeabilization time and RT reaction time were determined using the Visium Spatial Tissue Optimization Kit (10× Genomics). Second strand synthesis was subsequently performed on-slide at 65 °C for 15 minutes. All on-slide reactions were performed in a thermocycler with a metal slide-adapter plate. After second strand synthesis, samples were transferred to tubes for cDNA amplification and cleanup. Library quality was assessed using a Bioanalyzer High Sensitivity chip (Agilent 2100).

### Sequencing and data preprocessing

10× Genomics Visium libraries were pooled, denatured, and diluted, followed by paired-end sequencing on an Illumina Novaseq 6000. Sequencing data were processed using the Space Ranger pipeline v.1.3.1 (10× Genomics).

### Spatial transcriptome sequencing data analysis

Spatial RNAseq data of 11,016 barcoded spatial spots from three 10× Visium capture areas were analyzed using the Seurat V4.1.0 package. The spots with both nCount_Spatial and nFeature_Spatial larger than 500 were kept for further analysis. The feature counts were normalized and scaled by sctransform algorithm. The data from all three slices were merged and principal component analysis (PCA) was performed on a matrix composed of spots and gene expression (UMI) counts, reducing dimensions of the data according to the top 30 principal components. Uniform Manifold Approximation and Projection (UMAP) was used in this PCA space to visualize the data on reduced UMAP dimensions. The spots were clustered on PCA space using the Shared Nearest Neighbor (SNN) algorithm implemented as FindNeighbors and FindClusters with parameters k = 30, and resolution = 0.8. The method returned spot clusters representing anatomical regions in the tissues, which were then visualized on UMAP space using the SpatialDimPlot command. To accurately label anatomical regions, the spot clusters were compared with the brain map of Allen Brain Atlas. Then the markers were identified using FindAllMarkers function. The differentially expressed genes were calculated using DESeq2 at the cutoffs with *p* value < 0.05 and the absolute foldchange > 1.5. The spatial co-expression was calculated using Giotto (version 2.0.0.998) ^103^.

## Single-cell RNA sequencing

### Sample preparation and sequencing

The mice were anesthetized using 0.1 ml/10 g of 0.75% pentobarbital sodium solution, and perfusion was carried out to remove blood from the brain tissues. Two whole brains were taken from each genotype immediately after the mice were euthanized. The brain cell suspension mixtures were generated using MACS Adult Brain Dissociation Kit (Miltenyi Biotec, USA) following the manufacturer’s instruction. Cells were then processed using Chromium Single-cell 3’ v2 Library Kit (10× Genomics) (Oebiotech inc., Shanghai, China). Briefly, 21,000 cells were loaded onto a single channel of the 10× Chromium Controller. The mRNAs from approximately 19,000 cells were captured by nanoliter-sized gel beads, lysed within the emulsion, and reverse transcribed and barcoded using polyA primers with unique molecular identifier sequences before being pooled, amplified, and used for library preparation. The library was then sequenced in the Illumina Nova platform.

### SC-Seq data processing and analyses

The reads were mapped to the mouse genome reference sequences (mm10-1.2.0). The UMI count matrix was generated with cellranger’s (version 2.2.0) ‘cellranger count’ function for each library. To get high quality data, soupX was used to evaluate the contamination from ambient RNAs. The contamination fraction was set to 0.1 to remove the ambient RNAs. Then the object list was normalized with sctransform for integration by R toolkit Seurat (version 2.3.4). Dimensionality reduction of data was performed by principal component analysis (PCA) (N = 30 principal components) using R toolkit Seurat’s RunUMAP function, and the reduced data were visualized in two dimensions using UMAP. The resolution 0.2 was selected for the further analysis. The doublets were detected by R package DoubletFinder (version 2.0.3) and removed for the further analysis. The cell types were annotated based on the established annotation, using ConservedMarkers to call the most variable genes for each cluster, and enrichment analysis was performed on the cell markers obtained from literature ^67,68^ using DRscDB and clusterprofiler. The cutoffs for UMI count, gene number and mitochondrial ratio were set for each cell cluster (cell type) based on the actual data of the cell types respectively, to exclude the low-quality cells. The cutoffs were calculated as 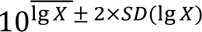, with X as UMI count, gene number or mitochondrial ratio respectively. We obtained 37,480 cells, 38,103 cells and 38,336 cells from the raw data of the wild-type, homozygous and heterozygous mice, respectively. After removing the low-quality cells, we obtained 29,877 cells, 30,037cells and 30,940 cells for the three genotypes, respectively. Differentially expressed genes were identified by FindMarkers function in the comparison for each cell type between the mutant and wild-type mice. Cell-cell communications were identified using CellChat (version 1.1.1).

### RNAscope

Frozen adult mouse brain tissue slices were used for RNA in situ hybridization using RNAscope® kit v2 (323100, Advanced Cell Diagnostics). The following mouse probes from Advanced Cell Diagnostics were used: Wnt4 (Cat# 401101-C4), Wnt5a (Cat# 316791-C3), and Foxj1 (Cat# 317091).

### Immunofluorescence

The antibodies used for immunofluorescence are DCN (AF1060, R&D), FZD4 (AF194, R&D), FZD6 (AF1526, R&D) and CTNNB1 (AF1329, R&D).

### Data availability

The authors are committed to the release of data and analysis results, with the understanding that rapid and transparent data sharing will benefit the scientific research community. Detailed analyzed data are included in supplementary excel tables. RNA-Seq, spatial RNA-seq data and SC RNA-Seq data are going to be deposited at GEO and will be available. All renewable reagents and detailed protocols will be available upon request.

### Code availability

The standard software packages used in data analysis are cited in the Methods.

